# Transdiagnostic Profiles of Behaviour and Communication Relate to Academic and Socioemotional Functioning and Neural White Matter Organisation

**DOI:** 10.1101/2021.11.29.21267002

**Authors:** Silvana Mareva, Danyal Akarca, The CALM team, Joni Holmes

**Affiliations:** Medical Research Council Cognition and Brain Sciences Unit, University of Cambridge 15 Chaucer Road, Cambridge, CB2 7EF; School of Psychology, Faculty of Social Sciences, University of East Anglia Norwich Research Park, Norwich, Norfolk, NR4 7TJ

## Abstract

**Background:** Behavioural and language difficulties co-occur in multiple neurodevelopmental conditions. Our understanding of these problems has arguably been slowed by an overreliance on study designs that compare deficit/diagnostic groups and fail to capture the overlap across different neurodevelopmental disorders and the heterogeneity within them.

**Methods:** We recruited a large transdiagnostic cohort of children with complex needs (*N* = 805) to identify distinct subgroups of children with common profiles of behavioural and language strengths and difficulties. We then investigated whether and how these data-driven groupings could be distinguished from a comparison sample (*N* = 158) on measures of academic and socioemotional functioning and patterns of global and local white matter connectome organisation. Academic skills were assessed via standardised measures of reading and maths. Socioemotional functioning was captured by the parent-rated version of the Strengths and Difficulties Questionnaire.

**Results:** We identified three distinct subgroups of children, each with different levels of difficulties in structural language, pragmatic communication, and hot and cool executive functions. All three subgroups struggled with academic and socioemotional skills relative to the comparison sample, potentially representing three alternative but related developmental pathways to difficulties in these areas. The children with the weakest language skills had the most widespread difficulties with learning, whereas those with more pronounced difficulties with hot executive skills experienced the most severe difficulties in the socioemotional domain. Each data-driven subgroup could be distinguished from the comparison sample based on both shared and subgroup-unique patterns of neural white matter organisation. Children with the most pronounced deficits in language, cool executive, or hot executive function were differentiated from the comparison sample by altered connectivity in predominately thalamocortical, temporal-parietal-occipital, and frontostriatal circuits, respectively.

**Conclusion:** These findings advance our understanding of commonly co-morbid behavioural and language problems and their relationship to behavioural outcomes and neurobiological substrates. Transdiagnostic Profiles of Behaviour and Communication Relate to Academic and Socioemotional Functioning and Neural White Matter Organisation

---

Difficulties with social communication, executive functions (EF), and behaviour are common across a range of neurodevelopmental conditions. Pragmatic or social communication impairments are included in the diagnostic criteria for autism spectrum disorders (ASD) and social (pragmatic) communication disorder. Difficulties with attention and/or behaviour are characteristic of attention deficit hyperactivity disorder (ADHD), and structural language problems are the hallmark of developmental language disorder (DLD) (American Psychiatric Association, 2013). While typically associated with separate disorders, these difficulties commonly co-occur across diagnoses. For example, children with ADHD often have pragmatic language difficulties (Green et al., 2014), and structural communication problems are common in both children with ADHD (Korrel et al., 2017) and ASD (Mandy et al., 2017). Similarly, behavioural and attentional difficulties occur in both ASD and DLD (Henry et al., 2012; Rommelse et al., 2011). These comorbidities suggest that problems with social communication, EF and behaviour might not be independent and disorder-specific. Instead, they likely share common aetiological origins and/or may interact dynamically, such that difficulties in one area might cascade developmentally to trigger problems in another (Masten et al., 2005).

Comorbidity is one of the main challenges to the current diagnostic system. This is further compounded by high levels of symptom variability within diagnostic categories: very different profiles of strengths and difficulties are common among children with the same diagnostic label (e.g. Astle et al., 2019; Kushki et al., 2019). For example, not all children with ADHD have EF impairments, and there is substantial heterogeneity among those who do (Kofler et al., 2019). This has led to widespread recognition of the limitations of diagnostic frameworks for guiding research and support strategies, and an increase in the application of transdiagnostic approaches for understanding neurodevelopment (see Astle et al., 2021 for a review).

## Transdiagnostic Approaches to Neurodevelopment

The dominant method for studying neurodevelopmental disorders involves comparing groups of children with a neurodevelopmental disorder to another diagnostic/deficit group, or a typically developing group, using univariate analytical approaches to tease apart differences. Transdiagnostic approaches adopt alternative recruitment and analytical strategies. In these studies, children with a variety of neurodevelopmental disorders are recruited, or diagnosis-based enrolment is replaced with enrolment based on needs. In the latter case, children who experience difficulties in the studied realm(s) are sampled regardless of whether their needs meet diagnostic thresholds (Holmes et al., 2019). Alternatively, large demographically representative cohorts, including individuals spanning the full spectrum of ability, are studied.

In terms of analyses, one goal is to identify dimensional characteristics that can predict or explain difficulties experienced across individuals irrespective of diagnostic status (e.g. Brislin et al., 2020). The relations between different dimensions and their links to neural/genetic factors can then be studied to identify potential mechanisms of shared or unique variance (Holmes et al., 2020; Parkes et al., 2021). A complementary analytical approach often described as *clustering* or *subtyping*^1^, focuses on deriving homogenous data-driven groupings of children with similar profiles of relative strengths and weaknesses along the studied dimensions (Feczko et al., 2018). These novel groupings can then be similarly used to facilitate the discovery of common aetiological pathways to specific difficulties, or to stratify individuals to support strategies appropriate for their needs.

Both approaches provide important and complementary insights into the aetiology of transdiagnostic symptoms and their implications for functioning. As one example, consider EFs. Dimensional approaches provide support for two dimensions of EF: cool EFs refer to the ability to regulate behaviour and cognition in emotionally neutral contexts, whereas hot EF skills are implicated in situations of stronger motivational and emotional valence (Castellanos et al., 2006). Studies using dimensional analyses reveal these dimensions of EF are related, but are associated with relatively distinct neural networks (Salehinejad et al., 2021) and have somewhat different predictive validity in terms of cool EF being more closely related to academic achievement and hot EF more predictive of disruptive behaviours (e.g. Willoughby et al., 2011).

Subgrouping approaches similarly reveal that different EF profiles have relatively distinct neural correlates and are associated with different outcomes. For example, Vaidya et al. (2020) identified three data-driven EF-subgroups across typically-developing, autistic and ADHD children. These subgroups explained more variance in frontal-parietal engagement (inferred via functional MRI) during a sustained attention task than diagnostic groupings. Similarly, Bathelt et al. (2018b) identified three distinct EF-subgroups in a transdiagnostic sample of struggling youth. Each subgroup was associated with different variations in white matter connections in prefrontal regions and there was more within-group homogeneity in the behavioural profiles of the derived subgroups than in groupings based on diagnosis. Both examples demonstrate that studying subgroups in transdiagnostic samples can facilitate the linking of behavioural phenotypes to neurophysiological mechanisms.

## Transdiagnostic Approaches to Neurodevelopmental Comorbidities

The examples of transdiagnostic designs described so far are typical of the field, with a focus on *one* domain of function (e.g., EF). An important next step is to apply these methods to multiple areas of functioning to advance our understanding of the co-occurrence of difficulties across *multiple* domains. We attempted to do this in an earlier study by exploring how symptoms of communication, behaviour, and EF difficulties relate to one another in a transdiagnostic cohort of children (Mareva et al., 2019). Using a network approach, we were able to investigate how specific symptoms cluster together and start to understand how these *clusters of difficulties* are linked.

Four densely interrelated clusters of symptoms were identified relating to structural language and learning, hot EF, pragmatic communication and peer relationships, and cool EF. The symptoms within each cluster did not align with the diagnostic features of any diagnosis. Moreover, specific symptoms bridged these clusters (e.g., inappropriate initiation of communication linked the pragmatic communication and hot EF clusters), providing some insight into the co-occurrence of symptoms across domains of difficulty. It may be the case that these bridging symptoms trigger difficulties through developmental cascades (Masten et al., 2005) or, alternatively, they may be particularly susceptible to the influence of symptoms in other domains. It is also possible that bridging symptoms do not signal interacting areas of difficulties, but instead reflect shared neural and/or genetic underpinnings.

## Linking Transdiagnostic Profiles to Aetiological Factors

Classifying individuals into heterogeneous and often overlapping diagnostic categories makes it difficult to understand the shared influence of aetiological factors (Kapur et al., 2012), but identifying groups of children with similar behavioural profiles, irrespective of diagnostic status, may provide one way to map symptoms on to underlying neurophysiology (e.g., Bathelt et al., 2018b; Vaidya et al., 2020). In the current study, we use this approach to identify groups of children with similar profiles across multiple domains of function and explore group differences in neural white matter organisation. Diffusion tensor imaging enabled us to estimate the macroscopic organisation of the white-matter connectivity of the brain *in vivo*. By analysing what are known as structural connectomes (Basser et al., 2000) we were able to explore how brain connectivity varied across children in the different subgroups. We focused on white matter because its maturation is an important aspect of post-natal neural development. White matter architecture enables efficient communication between discrete brain regions and shapes the processes underlying brain function (Honey et al., 2010). Variation in global white matter organisation has been linked to differences in general cognitive abilities, reading and mathematics skills (Bathelt et al., 2018a; Bathelt et al., 2019, Koenis et al., 2015). It has also been implicated in neurodevelopmental disorders such as ADHD and ASD (Beare et al., 2017; Cao et al., 2013; Qian et al., 2021), with both shared and distinct differences reported in white matter organisation across children with different neurodevelopmental disorders (Ameis et al., 2016; Qian et al., 2021). Examples of structural neural substrate variation implicated in multiple neurodevelopmental conditions include the corticostriatal circuits, thalamic radiations, and interhemispheric pathways (Ameis et al., 2016; Aoki et al., 2017; Arnsten & Rubia, 2012; Tung et al., 2021; Zhao et al., 2022).

## The current study

The aim of the current study was to identify homogenous subgroups of children with similar behavioural, EF, and communication strengths/difficulties in a large transdiagnostic sample. We then explored how the profiles of these transdiagnostic subgroups related to measures of white matter organisation, and academic and socioemotional functioning. Our previous study included a subsample of the currently studied population, and focussed on identifying *clusters of symptoms* that characterised the sample (Mareva et al., 2019). Individual differences at the symptom level can drive *clusters or subgroups of individuals* who share similar symptom profiles. Therefore, here, we use a complementary subgrouping approach to study how this space is occupied *by participants*, enabling us to further explore the associations between these profiles and academic and socioemotional functioning and underlying neurobiology. In other words, we searched for individuals with similar profiles across the four previously identified clusters of difficulties. The measures included in the subgrouping were chosen to capture transdiagnostic features (i.e., those that have been previously implicated in several disorders). These were measured by parent ratings on scales commonly used across health and educational settings. To explore the external validity of the subgroups and further characterise their profiles, we investigated how they differed on external (i.e., not included in the subgroup identification) measures of nonverbal cognitive ability, academic and socioemotional functioning, which are theorised to be related to interindividual differences in communication and EF (Arnold et al., 2020; Harpin et al., 2016; Helland & Helland, 2017). We did not formulate a hypothesis about the number of groups, their specific profiles, or the neural and external behavioural features that would differentiate them. Instead, we designed the study as an exploratory investigation aiming to address two broad questions: (1) can we identify robust subgroups of children presenting with distinct profiles of executive, language, and communication strengths/difficulties within a large transdiagnostic sample of struggling learners; (2) do such data-driven groups show any differences in academic attainment, socioemotional functioning, and white matter organisation relative to a comparison sample.

## Methods

The data presented here were collected as part of a cohort study at the Centre for Attention, Learning, and Memory (CALM). Data collection took part between 2014 and 2021.

### Recruitment

Full details about the CALM cohort are available in the study protocol (Holmes et al., 2019). Briefly, two groups of children were recruited: (1) a cohort of children aged 5 to 18 years who were referred by health and education practitioners for difficulties with attention, memory, and/or learning; (2) a comparison group who were not referred for difficulties. The latter group was recruited from the same schools attended by those who were referred via an open study invitation targeting children in the same age range. Children in both groups were enrolled into the study irrespective of diagnostic status or performance cut-offs, providing they met the following inclusion criteria: (1) native English speaker, no uncorrected sensory impairments, and (3) no confirmed presence of genetic or neurological conditions known to affect cognitive ability.

### Participants

The referred cohort included 805 children (69% male, *M _age_* = 9.48, *SD _age_* = 2.38) and the comparison 158 children (56% male, *M _age_* = 10, *SD _age_* = 2.33). The majority (63%) of the referred cohort were referred by education practitioners (e.g., educational psychologist, special educational needs coordinator); 33% were referred by health professionals (e.g., clinical psychologist, child psychiatrist); and 4% by speech and language therapists. Diagnoses were reported by the referrer and confirmed by parents. Most of the sample (60%) were undiagnosed, despite being recognised by a professional as having additional needs, and 8% had more than one diagnosis. Among those with diagnoses ADHD was the most common (*N* = 197), followed by ASD (*N* = 57), and diagnoses of learning disorders (e.g., dyslexia, dyscalculia, DLD, *N*=62). The non-referred sample consisted mostly of children without any diagnoses (96%), and among those with diagnoses one had ADHD, two had DLD, and two had dyslexia. For the comparison sample, diagnostic status was reported by parents/carers.

### Assessments

Each child completed a battery of neuropsychological assessments following the procedures documented in the assessment’s testing kits (see Holmes et al., 2019 for details). Parents/caregivers provided ratings of their child’s behaviour, communication, and socioemotional functioning. All children were invited to participate in an optional neuroimaging session within six months of their behavioural assessment (referred sample: *N* = 313, age at scan: *M* = 10.23, *SD* = 2.32; comparison sample: *N* = 77, age at scan: *M* = 10.75, *SD* = 2.01). The assessments included in the current study are described below.

#### Community Detection

##### Conners-3

The Conners Parent Rating Short Form 3^rd^ Edition (Conners, 2008) asks about the child’s ADHD-related difficulties in the past month. Item ratings are summarised into six subscales: Inattention, Hyperactivity/Impulsivity, Learning problems, Executive function, Aggression, and Peer relations. Raw scores were calculated, but age was regressed from each subscale to ensure the analyses were not biased by age.

##### Brief Rating Inventory of Executive Function (BRIEF)

The BRIEF (Gioia et al., 2000) captures behaviours related to EF. The 80 items in the checklist cover eight domains: Inhibition, Shifting, Emotional control, Initiation, Working memory, Planning, Organisation, and Monitoring. Raw scores were calculated, but age was regressed from each subscale to ensure the analyses were not biased by age.

##### Children’s Communication Checklist (CCC-2)

The CCC-2 (Bishop, 2003) focuses on communication strengths and weaknesses. Items are organised into ten subscales: Speech, Syntax, Semantics, Coherence; Inappropriate initiation, Stereotyped language, Use of context, Nonverbal communication, Social relations, and Interests. Raw scores were calculated, but age was regressed from each subscale to ensure the analyses were not biased by age.

#### Subgroup Validation

##### Strengths and Difficulties Questionnaire (SDQ)

The SDQ (Goodman, 1997) measures social and emotional functioning. Items are organised into five subscales (only four were analysed in the current study): Emotional Problems; Conduct Problems, Peer Problems, and Prosocial. The final SDQ subscale was omitted because it captures hyperactivity and overlaps with the Hyperactivity/Impulsivity subscale of the Conners that was included in the subtyping. Age-uncorrected SDQ scores were used in all analyses to allow for easier comparisons with other studies using this measure. Analyses using age-regressed scores supported the same conclusions (Table S2).

##### Reading and Mathematics

To assess children’s academic abilities the Word Reading and Numerical Operations subtests of the Wechsler Individual Achievement Test-II (Wechsler, 2005) were administered. Raw scores for each subtest were converted to age-referenced standard scores to enable performance to be compared to age-expected norms.

##### Nonverbal cognitive ability

The Matrix Reasoning subtest of the Wechsler Abbreviated Scale of Intelligence (Wechsler, 2011) was administered as an index of non-verbal cognitive ability. The raw score, corresponding to the number of correctly completed matrices, was converted into an age-referenced *T*-score, allowing for straightforward interpretation relative to age-expected levels.

### MRI Data Acquisition

The magnetic resonance imaging (MRI) data were collected on a Siemens 3 T Prisma-fit system using a 32-channel quadrature head coil. T1-weighted volume scans were acquired using a whole-brain coverage 3D Magnetization Prepared Rapid Acquisition Gradient Echo (MP RAGE) sequence acquired using 1 mm isometric image resolution. Echo time was 2.98ms, and repetition time was 2,250ms. Diffusion scans were obtained using echo-planar diffusion-weighted images with an isotropic set of 68 noncollinear directions, using a weighting factor of *b* = 1,000s × mm^−2,^ interleaved with 4 T2-weighted (*b* = 0) volume. Whole brain coverage was based on 60 contiguous axial slices and isometric image resolution of 2 mm. Echo time was 90ms and repetition time was 8500ms. Both MRI pre-processing and reconstruction were performed using QSIPrep 0.13.0RC1, which is based on Nipype 1.6.0 (Gorgolewski et al., 2011). All pre-processing steps are reported in the Supplementary Materials. Whole-brain white matter connectivity matrices (i.e. connectomes) were constructed for each child based on the Brainnetome atlas (Fan et al., 2016). For each pairwise combination of regions (*N* = 246), the number of streamlines intersecting them was estimated and transformed to a 246 x 246 streamline matrix.

### Statistical Analysis Overview

A data-driven community detection algorithm was applied to the child-by-child associations across the behaviour, EF, and communication ratings in the referred sample to identify subgroups of children with similar profiles of behavioural and communication strengths and difficulties. The demographic and diagnostic characteristics, academic performance, socioemotional ratings, and neural white matter connections were then compared across the derived subgroups and the comparison sample. The details for each step of the analysis are presented below.

#### Community Detection

A network was built to represent the child-by-child correlations across the 24 subscales of the CCC-2, Conners-3, and BRIEF questionnaires for those in the referred sample. Child-by-child correlations were the focus of the community detection to capture similarities and differences in children’s profiles across the subscales. The method has been previously used to identify subgroups of children with similar neuropsychological profiles, temperaments, and EF-related behavioural problems (Bathelt et al., 2018b; Fair et al., 2012; Karalunas et al., 2014). Before estimating the child-by-child correlations, missing data was estimated and age was regressed from each scale to ensure the community detection was not biased by age (see Supplementary Materials for details). The community detection was based on the Louvain weight-conserving algorithm (gamma set at default value of 1), as implemented in the Brain Connectivity Toolbox (BCT: Rubinov & Sporns, 2010). The strength of the community separation was quantified using the asymmetric modularity (*Q*) index (Rubinov & Sporns, 2011), which conceptually represents the overall segregation between identified communities, with values above 0.3 considered evidence for sufficient community separation (Newman & Girvan, 2004, Blondel et al., 2008). This method performs well in recovering the true number of communities across a range of conditions and simulations suggest the sample size of the referred cohort was suitable for this analysis (Agelink van Rentergem et al., 2022; Gates et al., 2016). To reach a stable community assignment, we applied the consensus clustering method with 1000 iterations (Lancichinetti & Fortunato, 2012), a procedure that has demonstrated robustness even at substantial levels of noise in the data (Bathelt et al., 2018b).

#### Subgroup Profiles and Behavioural Validation

A series of two-tailed *t*-tests with Holm corrected *p*-values were run to compare the profiles of the derived subgroups to each other, and to the comparison group, to quantify variation in the severity of difficulties across data-driven groups and to capture differences relative to the non-referred children. To facilitate the interpretation of the subgroup profiles, the communication and behavioural data used in the community detection was reduced via Principal Component analyses (PCA) and component scores were extracted and compared across the groups. Data from both the referred and comparison samples was combined for the PCA, and the optimal number of components to retain was chosen based on the results of parallel analyses (see Supplement). Separate PCAs were run for the measure of communication (CC2) and the two measures of behaviour combined (BRIEF and Conners). Additional group comparisons were conducted on measures not included in the community detection to externally validate and further characterise the profiles of the subgroups.

#### Neuroimaging Analysis

Neural white matter connectome data was analysed using graph theory. The nodes of the network were the 246 regions of the Brainnetome atlas (Fan et al., 2016) and the connections (or edges) corresponded to the number of streamlines intersecting each pair of regions. All connectome analyses are based on weighted unthresholded matrices. This choice was motivated by observing that the application of consistency-thresholding (Roberts et al., 2017) to retain the top 30% or 20% of most consistent edges (on the assumption that the connections with the highest inter-subject variability are spurious) fully reproduced the results based on unthresholded matrices. We explored whether and how each derived subgroup differed from the comparison sample and did not compare the data-driven subgroups to one another. This approach was chosen to reduce the number of comparisons and to investigate both common and subgroup-unique differences among the referred children relative to the non-referred children. Due to the presence of outliers in the data, the global and local metrics of the data-driven subgroups were compared to the comparison group using 10% trimmed-means pairwise *t*-tests with false-discovery rate correction.

##### Global and Local Metrics

At the whole-brain level, we focussed on global efficiency and global clustering coefficients. These metrics were chosen because they have been previously linked to children’s educational attainment and general cognitive abilities (Bathelt et al., 2018a; Bathelt et al., 2019; Koenis et al., 2015) and have been implicated in neurodevelopmental disorders such as ADHD (Cao et al., 2013). Global efficiency describes the potential for information exchange in the network and is the average inverse distance from any node (i.e. brain region) to any other node (Sporns et al., 2007). Global clustering quantifies the fraction of each node’s neighbours that are also neighbours of each other. The global clustering metric was normalised according to the average of 1000 random graphs using the algorithms for weighted undirected networks implemented in the GRETNA toolbox (Wang et al., 2015). Subsequently, to explore the local properties of the connectomes with the minimal number of comparisons, the 246 brain regions in the Brainnetome atlas (Fan et al., 2016) were grouped according to their corresponding intrinsic connectivity networks (ICN) as defined by Yeo et al. (2011): Default mode, Dorsal attention, Frontoparietal, Limbic, Somatomotor, and Visual networks. Subcortical regions were also grouped together. The sum of all connections within each network (and the subcortex) of the connectome was calculated. This approach is based on the known structural and functional organisation properties of the brain and was therefore favoured as a balanced alternative to hypothesis-driven pre-specified regions of interest approach or a fully data-driven reduction of the connectomes. For each data-driven subgroup, networks that showed a significant difference from the comparison group were selected for further analyses. Subsequently, the node strength (i.e., the sum of all connections) in each region within the networks flagged as significantly different was tested against the comparison group. The functional characterisation of these regions was based on the behavioural domain metadata labels of the BrainMap Database (www.brainmap.org/taxonomy), which uses both forward and reverse inferences (Eickhoff & Grefkes, 2011; Fox et al., 2014). Prior to the analyses, age, gender, and average frame displacement were regressed from each graph metric using a robust regression approach to account for outliers. Controlling for these covariates is recommended and is standard practice in neurodevelopmental neuroimaging research (Alexander-Bloch et al., 2016; Jones et al., 2021; Luna et al., 2021).

##### Network-Based Statistics (NBS)

To investigate differences in the connections between regions we used NBS, which in many cases offers greater statistical power compared to traditional correction methods (Zalesky et al., 2010). This is a whole-brain data-driven method and is described in detail in the Supplement. Briefly, each data-driven subgroup was compared to the non-referred group to identify subnetworks that differentiate the groups. The following parameters were used: *t*-threshold 2.8, intensity measure of size, and family-wise error (FWE) *p* < .01. Age, gender, and average frame displacement were used as covariates in all analyses. To ensure the robustness of the results, we repeated all procedures by varying the target *t*-threshold, exploring the extent-based measure of size, and setting a more liberal FWE-correction (*p* < .05).

## Results

### Community Detection

Community detection applied to the child-by-child correlation network based on the Louvain algorithm identified three subgroups of children (*Q* = .43; Figure S1). The value of the *Q*-metric was above 0.3, suggesting a good level of community separation (Blondel et al., 2008).

### Subgroup Profiles

The profiles of the subgroups identified by the community detection and the comparison group across the 24 subscales are displayed in Figure 1a. Descriptive statistics and Cohen’s *d* effect size are presented in Table S1. To aid the interpretation of the differences across the groups, data was reduced with two PCAs, one applied to the CCC-2 and a second to the combined BRIEF and Conners data (see Figure S2-3 for correlations). This decision allowed us to consider communication and behaviour separately. The CCC-2 data reduction identified two components capturing structural and pragmatic communication, which in combination explained 80% of the variance. Two components broadly corresponding to hot EF and cool EF summarised the Conners and BRIEF data, cumulatively explaining 68% of the variance (Figures 1b and 1c; full report in Supplement). As a sensitivity analysis, in the supplement, we also report the results of parallel analysis and PCA applied to the CCC-2, Conners, and BRIEF data together. This approach supported broadly similar conclusions, with the exception that hot EF and pragmatic skills loaded on a single component. The full details are available in the supplement (Figure S6 and S7).

**Figure 1.**
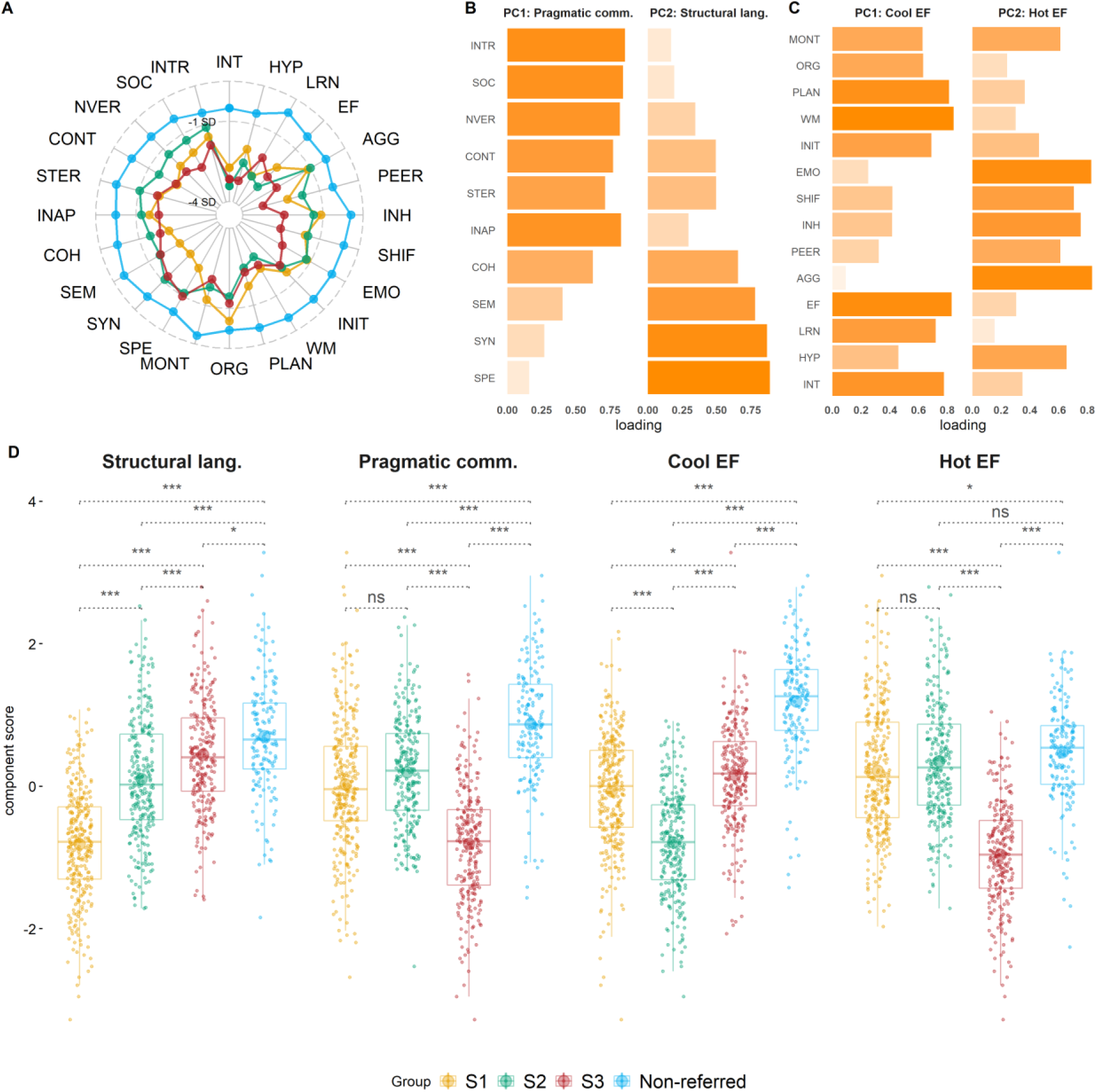
Profiles of the data-driven subgroups and the comparison group. *Note.* **Panel A** shows group profiles across all BRIEF, Conners, and CCC-2 subscales used in community the detection. To put all measures on the same scale, age-referenced subscale scores were converted to z-scores where higher values indicate strengths and lower values indicate difficulties. **Panel B** shows the varimax loadings from Principal component analyses (PCA) of the CCC-2 subscales based on combined data from the referred and comparison samples. The first principal component (PC1) explained 46% of the variance and was labelled Pragmatic communication, the second principal component (PC2) explained additional 34% of the variance and was labelled Structural language. **Panel C** shows the loadings from the same analyses applied to the BRIEF and Conners data. PC1 explained 37% of the variance and was labelled Cool EF and PC2 explained additional 31% of the variance and was labelled Hot EF. **Panel D** shows group performance across the four components identified in the PCA: higher values indicate strengths and lower values indicate difficulties. Comparisons are based on two-tailed *t-*tests, *p*-values are Holm-corrected. *** p <.001; ** p < .01, * p < .05. S1 = Subgroup 1; S2 = Subgroup 2; S3 = Subgroup 3; Conners (Conners Parent Rating Short Form 3rd Edition) subscales; EF = Executive function; INT = Inattention; HYP = Hyperactivity/Impulsivity; LRN = Learning Problems; AGG = Aggression; PEER = Peer Relationships; BRIEF (Brief Rating Inventory of Executive Function) subscales: INH = Inhibition; SHIF =Shifting; EMO = Emotional Control; INIT = Initiation; WM = Working memory; PLAN = Planning/Organisation; ORG = Organisation of Materials; MONT = Monitoring; CCC-2 (Children’s Communication Checklist) subscales: SYN = Syntax; SEM = Semantics; COH = Coherence; INAP = Inappropriate Initiation; STER = Stereotyped Language; CONT = Use of Context; NVER= Nonverbal Communication; SOC = Social Relations; INTR = Interests.

**Table 1.**
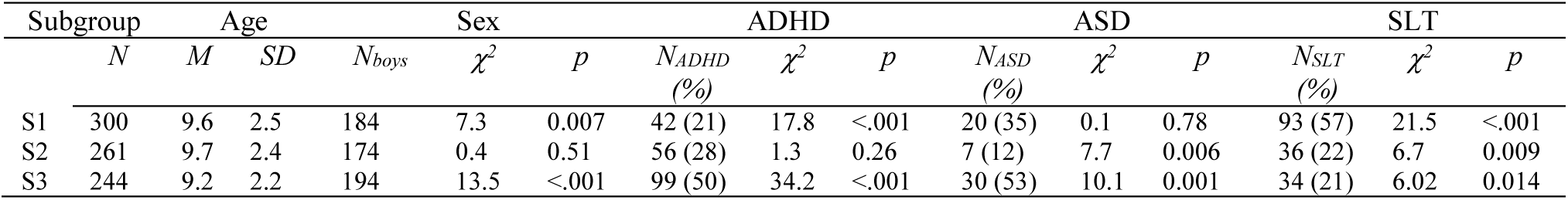
Subgroup demographics. *Note*. ADHD = Attention-deficit hyperactivity disorder; ASD = Autism spectrum disorder; SLT = Speech and Language therapy in the last two years or ongoing. S1 = Subgroup 1 (most severe structural language difficulties); S2 = Subgroup 2 (most severe *cool* executive difficulties); S3 = Subgroup 3 (most severe difficulties with *hot* executive skills & pragmatic communication).

Group profiles across the four components are displayed in Figure 1d. The data-driven subgroups had significantly more difficulties across all components relative to the comparison sample, except for Subgroup 2 (S2) who had similar hot EF to the comparison sample. The children in each subgroup had different patterns of specific strengths and difficulties. Subgroup 1 (S1) were characterised by poor structural language, children in S2 had weaknesses in cool EF, and children in Subgroup 3 (S3) had pronounced difficulties with hot EF and pragmatics (Table S2). The same patterns were evident in the subgroup profiles on the subscales that most strongly loaded on the respective principal components (Table S1 and Figure 1a). One-way ANOVA suggested no significant differences in age across the subgroups (*F* (1,803) = 3.138, *p* = .08, Table 1), but pairwise *t*-tests with Holm correction for multiple comparisons suggested that children in S2 were significantly older than children in S3 (*t* = 2.56, *p _corrected_* = .03), with an average difference of 6.34 months. Using a chi-square test, which compared the overall proportion of boys and girls in the whole referred sample to the proportion observed in each subgroup, we observed that girls were overrepresented in S1, while boys were overrepresented in S3 (Table 1). The same chi-square analyses were used to investigate whether there were differences in the prevalence of different diagnoses across subgroups. We focused on children with ADHD and ASD because overall numbers for other diagnoses were too low to make meaningful comparisons. Children with ADHD and ASD were represented in each subgroup, but children with ADHD were overrepresented in S3 and underrepresented in S1, and children with ASD were underrepresented in S2 and overrepresented in S3 (Table 1). Analyses comparing the prevalence of ADHD subtypes across clusters were not pursued due to most children with ADHD having a diagnosis of Combined type ADHD. Only one child had a diagnosis of predominantly Hyperactive/ Impulsive ADHD (included in S3) and only fourteen a diagnosis of the predominantly Inattentive type (two cases included in S1, seven in S2, and five in S3). Finally, children who had recently received Speech and Language Therapy were overrepresented in S1 and underrepresented in the other two subgroups (Table 1).

### Subgroup Profiles: Behavioural Validation

The groups were compared on external measures not used in the community detection (Figure 2). In terms of socioemotional functioning and cognitive and academic performance, all three subgroups derived from the referred sample had significantly more difficulties than the comparison sample (Figure 2). There were differences in the patterns of severity among the subgroups. Children in S1 had the most pronounced difficulties with maths, reading, and non-verbal reasoning, scoring significantly lower than the other two data-driven groups. Children in S3 had better performance than the other two subgroups in math and reading, but significantly more difficulties with emotion, conduct, peer relations, and prosocial behaviour (Figure 2).

**Figure 2.**
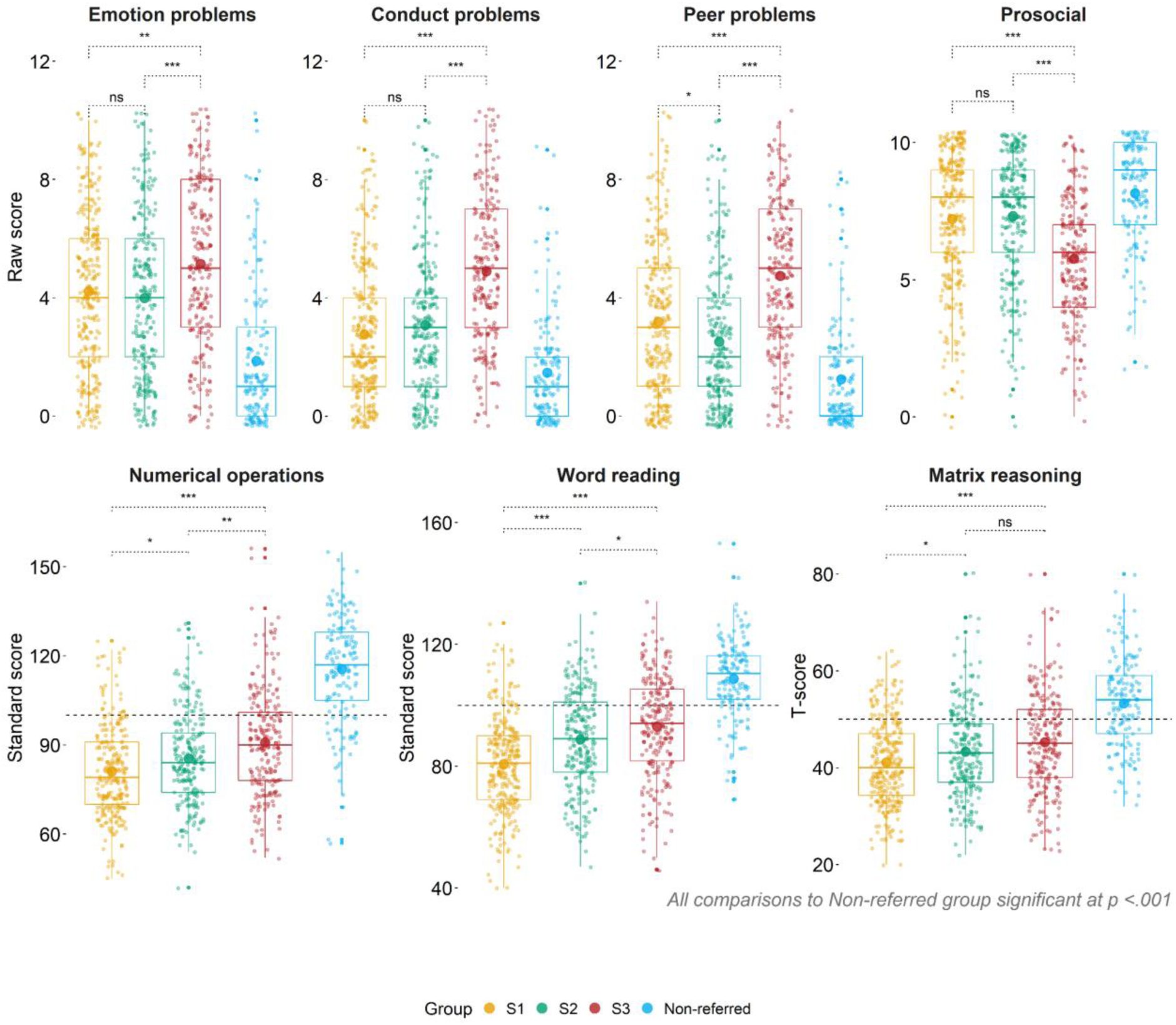
Group comparisons across measures of socioemotional functioning, cognitive, and academic skills. *Note.* The top panel includes the Emotion problems, Conduct problems, Peer problems, and Prosocial subscales derived from the Strengths and difficulties questionnaire (SDQ, parent-report). Higher scores indicate more difficulties, except the Prosocial scale, which has the reverse interpretation. The bottom panel shows performance on the Word reading and Numerical operations subsets of the Wechsler Individual Achievement Test-II, and the Matrix reasoning subset of Wechsler Abbreviated Scale of Intelligence II. All scores are based on the age-referenced norms: higher scores indicate better performance. The grey-dotted line represents the age-expected mean. For both panels, the observed mean in each group is represented by the large dot within each boxplot. All comparisons are based on two-tailed *t*-tests, *p*-values are Holm-corrected. S1 = Subgroup 1 (most severe structural language difficulties); S2 = Subgroup 2 (most severe *cool* executive difficulties); S3 = Subgroup 3 (most severe difficulties with *hot* executive skills & pragmatic communication). *** *p* <.001; ** *p* < .01, * *p* < .05

Overall, three subgroups were identified capturing children with principal difficulties with structural communication (S1), cool EFs (S2), and hot EFs and pragmatics (S3). Considering measures, which were not included in the community detection, there was a consistent pattern in which children in S3 had more pronounced socioemotional difficulties, whereas children in S1 and S2 showed more difficulties with cognitive and academic skills.

### Subgroup Profiles: Neural White Matter

#### Graph Measures

At the global level, S1 and S2 had significantly lower global efficiency relative to the comparison sample (S1, *N*_imaging_ =110: *p_corrected_* = .04; S2, *N*_imaging_ = 121: *p*_corrected_ = .04). The difference between the comparison sample and S3 (*N*_imaging_ = 82) was not significant (*p_corrected_* = .10). There were no differences in global clustering between the comparison sample and any of the subgroups (uncorrected *p*-s > 0.25). These results are presented in Figure S8. At the ICN-level, all subgroups had weaker overall connections in the limbic network and subcortical areas relative to the comparison sample (Figure S9). Follow-up analyses showed that relative to the non-referred sample all subgroups had reduced regional strength in the same sub-regions of the basal ganglia (BG), hippocampus, thalamus, inferior temporal gyrus (ITG), and fusiform gyrus (FFG). Some unique patterns of reduced regional strength were observed for each subgroup relative to the comparison group (Figure 3 and Tables S3 and S4). For children in S1, unique differences were observed in sub-regions of the BG and ITG, which the BrainMap taxonomy labels as primarily implicated in cognition, emotion, action execution, memory, and semantics. For children in S2, unique differences were within the parahippocampal gyrus (PHG), superior temporal gyrus (STG), ITG, BG, and amygdala, areas that have previously been implicated in cognition, emotion, social cognition, and perception. Finally, for children in S3, these differences were in the BG, cingulate gyrus, and thalamus, which are linked to action execution, perception, somesthesis, and cognition (Figure 3 and Tables S3 and S4).

**Figure 3.**
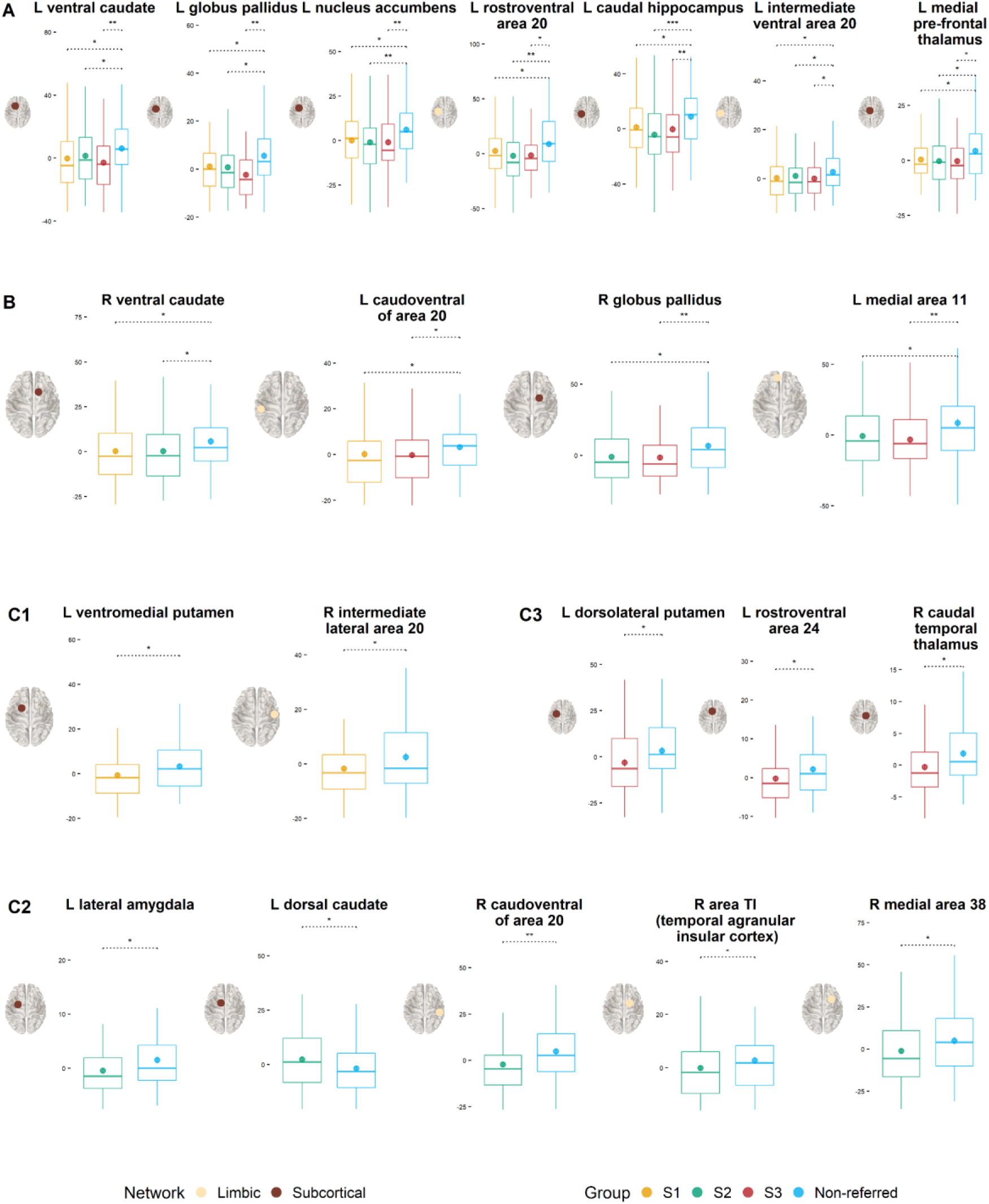
Comparison of the regional strength of connections across groups. *Note*. **Panel A** shows regions within the subcortex and the limbic network that showed reduced connection strength in all data-driven subgroups relative to the comparison group. **Panel B** shows subcortical and limbic regions that had reduced connection strength in two subgroups relative to the comparison group. Note that non-significant comparisons are omitted from the figure. **Panel C1** shows subcortical and limbic regions that were significantly different between S1 and the comparison group. **Panel C2** shows subcortical and limbic regions that were significantly different between S2 and the comparison group. **Panel C3** shows subcortical and limbic regions that were significantly different between S3 and the comparison group. S1 = Subgroup 1 (most severe structural language difficulties); S2 = Subgroup 2 (most severe *cool* executive difficulties); S3 = Subgroup 3 (most severe difficulties with *hot* executive skills & pragmatic communication). See Tables S3 and S4 in supplement for descriptive statistics, *p*-values, and effect sizes.

#### NBS

Subnetworks that were significantly different for each subgroup relative to the comparison group are displayed in Figure 4. There was greater connectivity in the non-referred group relative to S1 in a subnetwork comprising five connections across limbic—subcortical and subcortical—subcortical areas (*p_corrected_* = .001). This subnetwork involved six regions located within the BG, orbital gyrus, and thalamus (Table S5). For S2, there was reduced connectivity relative to the comparison group in a subnetwork spanning nine connections across ten regions (*p*_corrected_ = .003). This subnetwork included connections across the following networks: somatomotor — dorsal attention, limbic — dorsal attention, dorsal attention — visual, frontoparietal — somatomotor, visual — visual, and dorsal attention — subcortical. The involved regions fell within the lateral occipital cortex (LOC), medioventral occipital cortex, orbital gyrus, PHG, precuneus, superior parietal lobule, STG, and thalamus (Table S6). Finally, S3 had reduced connection strength relative to the comparison group in a subnetwork comprised of eight links (*p_corrected_* = .002) across limbic — subcortical, limbic — visual, visual — subcortical, and subcortical — subcortical regions. The eight regions involved formed part of the BG, LOC, middle frontal gyrus, orbital gyrus, precuneus (this region was also part of the network that differentiated S2), and thalamus (Table S7). Sensitivity analyses supported the robustness of the findings: taking the extent-based measure of size fully reproduced the results; and setting the *t*-threshold in the range of 2.6 to 3, produced similar results, which followed the expected tendency to discover larger subnetworks at lower *t*-thresholds (Figure S11; for an investigation of the influence of the *t*-threshold see Beare et al., 2017). Results based on the more liberal FWE-corrected threshold of *p* < .05 suggested further differences for S3 (Figure S10).

**Figure 4.**
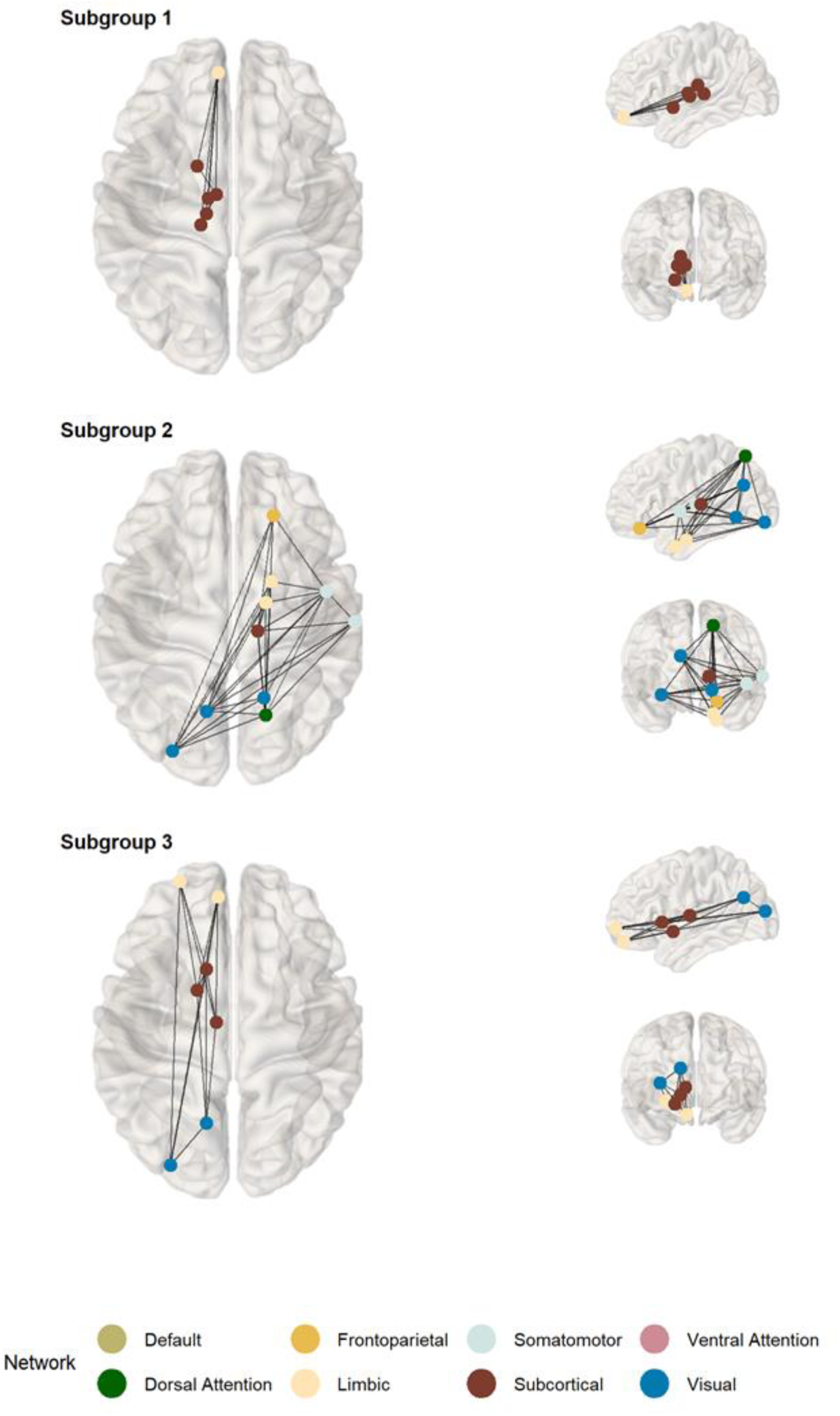
Subnetworks of the neural white matter connectome identified as significantly weaker in each data-driven subgroup relative to the comparison group. *Note.* Subgroup 1: most severe structural language difficulties; Subgroup 2: most severe *cool* executive difficulties; Subgroup 3: most severe difficulties with *hot* executive skills & pragmatic communication. Nodal labels in Tables S5-S7.

Overall, the three subgroups showed a mix of similar and subgroup-unique patterns of differentiation from the comparison sample. At the global level, the two groups (S1 and S2) with the most pronounced difficulties with academic achievement showed reduced global efficiency compared to the non-referred group. At the ICN-level, all subgroups showed reduced connection strengths in the limbic network and the subcortex relative to the comparison sample. There were both common and subgroup-specific differences in the connection strength of regions within the limbic network and the subcortex. Finally, at a whole-connectome level, we identified subnetworks that differentiated each subgroup from the comparison sample.

## Discussion

The current study adopted a data-driven approach to identify subgroups of children with homogeneous profiles across different domains of function as measured by ratings of communication, behaviour, and EF. A transdiagnostic approach was used: enrolment was based on cognitive and academic needs rather than diagnostic status. Differences in brain structure and behaviour were compared across the subgroups in relation to a comparison group. Three subgroups of children were identified. Each performed more poorly than the comparison group across measures of communication, behaviour, and EF, and was distinguished from the other subgroups by different profiles of strengths and weaknesses in these areas. These differences extended to measures of cognitive, academic, and socioemotional functioning that were not included in the identification of the subgroups. Shared and specific patterns of differences in neural white matter organisation were observed across the groups. These results are discussed below.

### Subgroups

Three data-driven subgroups were identified based on parent/carer ratings of behaviour, communication, and EF in a large transdiagnostic sample of children referred by practitioners for difficulties in attention, learning, and/or memory. One subgroup (S1) was characterised by relative difficulties in structural language use, a second by *cool* cognitive difficulties (S2), and a third by co-occurring pragmatic communication difficulties and *hot* affective cognitive problems (S3). All three subgroups had greater difficulties in behaviour, communication, EF, socioemotional functioning, and academic attainment relative to a demographically matched comparison group of non-referred children.

There were both similarities and differences in the way neural white matter was organised in the three subgroups relative to the comparison group. The two subgroups with the most pronounced difficulties in cognitive and academic skills (S1 & S2) showed reduced global efficiency, which has been previously linked to educational attainment (e.g., Bathelt et al., 2019; Lou et al., 2019). However, no differences were observed in global clustering coefficients. At the ICN-level, all groups showed reduced connectivity within the subcortex and the limbic network relative to the comparison group. Exploring differences in the connectivity of specific regions within these areas, all three subgroups showed reduced regional strength relative to the comparison sample in subregions of the hippocampus, BG, ITG, thalamus, and FFG, which the BrainMap taxonomy labels as being involved in cognition, memory, action execution, language, interoception, and emotion. These findings demonstrate that the subgroups of referred children identified by the data-driven approach have shared and distinct behavioural and neural features relative to a non-referred comparison group. In the following sections, the detailed profiles of the three subgroups are considered.

### S1

The children in S1 were characterised by elevated difficulties with structural language skills relative to the other subgroups, and those who had attended Speech and Language Therapy in the past two years were overrepresented in this group. They also had the most severe difficulties in learning. Finding concurrent difficulties in structural language use and academic achievement is consistent with previous reports (Dockrell et al., 2011; Mareva & Holmes, 2019), and might reflect underlying phonological processing difficulties that have been implicated in language, reading, and maths difficulties (e.g., Amland et al., 2021).

The neural subregions that showed significantly reduced connectivity only for children in S1 relative to the comparison group were in the right ITG and the left ventromedial putamen. A left-lateralised subnetwork of primarily limbic-subcortical connections that involved subregions of medial orbitofrontal cortex, thalamus, and BG also distinguished this group. Consistent with their language and learning difficulties, corticostriatal and thalamocortical pathways are involved in procedural learning, language development, goal-directed behaviour, and reward processing (Arnsten & Rubia, 2012; Krishnan et al., 2016). Furthermore, most connections in the identified subnetwork involved left medial area 11, which the BrainMap taxonomy labels as functionally implicated in language and orthography.

### S2

The most pronounced area of weakness for the second subgroup (S2) was in cool EF where their scores were lower than those of children in the other two subgroups. These difficulties encompassed everyday difficulties in attention, planning, and working memory - skills that have been implicated in classroom learning (Peng et al., 2018). Consistent with this, children in this subgroup had difficulties in maths and reading, although these were not as severe as those for children in S1. Relative to the other two subgroups, children in S2 had relative strengths in social skills and affective cognition, with comparable hot EF ratings to the comparison group.

Subregions that uniquely deviated in S2 included regions within caudate, PHG, ITG, STG, and lateral amydgala. The subnetwork that differentiated them from the comparison group was relatively widespread and involved right-lateralised temporal-parietal pathways and medioventral and lateral occipital regions. Some of the implicated subregions of these dorsal attention and visual networks are known to interact to suppress attention to irrelevant stimuli (Castellanos & Proal, 2012; Shulman et al., 2009), and as such reduced connectivity in this neural circuit might contribute to the cool EF-related difficulties characteristic of this subgroup. In particular, seven of the nine connections within the identified subnetwork involved right caudal area 7, which is implicated in cool EFs such as attention, working memory, inhibition, and spatial cognition (for details see http://atlas.brainnetome.org/, Fan et al., 2016).

### S3

The children in S3 were characterised by having the most severe difficulties with hot EF and pragmatic communication, but relative strengths in structural language skills, maths and reading, compared to the children in the other two subgroups. Strong associations between social communication skills and affective behavioural problems have been reported previously (e.g., Hawkins et al., 2016; Mareva et al., 2019), and difficulties in these areas commonly co-occur in ADHD and ASD (Green et al., 2014) alongside socioemotional difficulties (Staikova et al., 2013). Indeed, a disproportionate number of autistic children and children with an ADHD diagnosis were assigned to this subgroup, and they had substantial problems with behavioural conduct, emotion, peer relationships, and prosocial behaviour. Identifying a subgroup with this profile and composition suggests the intersection of pragmatic communication, hot EF, and socioemotional difficulties may be relevant for understanding some of the comorbidity between ADHD and ASD.

The neural characteristics that specifically differentiated children in S3 from the comparison group were regions within the putamen, thamalus, and cingulate gyrus. They had reduced connectivity strength in a left-lateralised, primarily frontostriatal subnetwork, which also included regions of the visual network (precuneus and LOC). Consistent with the severity of their hot EF difficulties, these circuits play a role in goal-directed behaviours related to reward, affect, and motivation (Arnsten & Rubia, 2012).

### Summary of Profiles

In summary, we identified three subgroups of children with distinct communication, behavioural, and EF profiles. These subgroups were characterised by primary difficulties in structural language, cool EF, or hot EF and pragmatics, respectively, and provide initial evidence for three alternative but related pathways to academic and socioemotional difficulties. While a greater number of children receiving speech and language therapy were assigned to the structural language subgroup, and more autistic children and those with ADHD to the hot EF and pragmatics subgroup, none of the subgroup profiles aligned with the diagnostic features of a particular disorder, and children with each of these diagnoses were present in each of the three subgroups. This finding suggests the subgroups were not synonymous with disorder-based categories, adding to growing support for transdiagnostic approaches to understanding neurodevelopment (Astle et al., 2021).

The three subgroups were further distinguished by patterns of differences in the connectivity of circuits previously implicated in language, executive and visual attention, and reward processing. These differences partially correspond to previously reported neurobiological correlates of the behavioural difficulties of the subgroups, suggesting they may be distinguished at the neural level. That said, not all regions that uniquely differentiated the subgroups from the non-referred sample had clear links to their behavioural profiles. It should also be noted that all subgroups had reduced connection strength within the limbic network and the subcortex, relative to the comparison group, and shared several atypicalities within the same subcortical and temporal subregions. Furthermore, their profiles of behavioural weaknesses were all relative, meaning the correspondence between brain and behaviour was not one-to-one.

### Theoretical and Practical Implications

Following decades of research shaped by diagnostic categories, the field of neurodevelopmental difficulties is currently undergoing a transition in which exploratory research into how children and their characteristics are clustered is taking priority (Astle et al., 2021). Such research is key for testing, and where necessary accordingly modifying, predominant assumptions about diagnostic boundaries or common factors that explain the associations between symptoms (Astle & Fletcher-Watson, 2020). While the current study was not designed to explicitly test or falsify a given theory, the finding that children with different diagnoses were represented in all data-driven clusters challenges “core deficit” theories that assume that a single mechanistic impairment can explain the profile of a particular diagnostic group. Instead, our results are more consistent with accounts emphasising the possibility that multiple causal pathways can lead to the same behavioural phenotype (Cicchetti & Rogosch, 1996). Relatedly, we found little evidence for neat or direct mappings between isolated neural structures and behavioural phenotypes. Most brain atypicalities associated with a given behavioural phenotype were also observed across the other phenotypes, casting doubt on claims that focal neural deficits underlie a given neurodevelopmental disorder. Our findings are instead consistent with the predictions of neuro-constructivist theories that assume developmental difficulties have widespread effects that result from a brain that has developed differently over a number of years (Johnson, 2011; Karmiloff-Smith, 2009).

The categorisation of neurodevelopmental difficulties into discrete disorders has practical merits in providing health and education practitioners with a pragmatic system for selecting and allocating support. It also provides many young people with a sense of identity. Receiving a diagnosis can prove a pivotal moment in someone’s life, enabling them to identify with their community. Yet our data, and that of others, highlight that current diagnostic approaches do not capture what it is like to have additional needs: the clusters of behavioural symptoms that children experience do not map on the diagnostic criteria currently used to identify and support children’s needs. As such, a more flexible, child-centred approach is needed in which intervention decisions are based on individual needs and not primary diagnoses (Finlay-Jones et al., 2019). Attempts to integrate the neurodevelopmental transdiagnostic framework into clinical settings are just emerging, and in due course we will start to gain insights into their efficacy and feasibility (Boulton et al., 2021). For now, we advocate transdiagnostic approaches, which align with the neurodiversity paradigm (Fletcher-Watson, 2022), as a means of promoting more inclusive research and practice.

### Limitations & Future directions

There were several caveats to the current subtyping approach. First, the community detection was based on parent ratings, which are prone to subjective bias. However, the differences between the subgroups identified through these ratings were reflected in differences in performance-based measures of cognition and learning, providing some validity to the ratings and suggesting the algorithm was not overfitting the data. Second, the wide age range and cross-sectional nature of the cohort did not allow us to explore questions about age-related heterogeneity or developmental continuity. Nonetheless, we did observe a significant age difference between S2 and S3, suggesting the S3 phenotype which was associated with greater social and emotional problems may be more prevalent in younger children. Finally, despite the sample being substantially larger than that typically used in the developmental neuroimaging literature, sample size considerations did not allow us to directly compare the neural profiles of the identified subgroups to one another. Equally important differences in white matter organisation across subgroups might exist, but larger datasets would be needed to identify them.

## Conclusion

This study demonstrates the value of data-driven subgrouping approaches for understanding common, complex and co-occurring neurodevelopmental difficulties across multiple domains and their relationships to behavioural outcomes and neurobiology. It shows that homogeneous groups can be identified and differentiated in terms of distinct profiles of relative strengths and difficulties across communication, executive function, and behaviour. The identified subgroups provide initial evidence for three alternative but related developmental pathways to difficulties with academic and socioemotional functioning.

## Data Availability

The CALM dataset is not yet available as the study is still ongoing. The data will be made available via managed open access once the study is complete

## Acknowledgements

The CALM Team includes lead investigators Duncan Astle, Kate Baker, Susan Gathercole, Joni Holmes, Rogier Kievit and Tom Manly. Data collection is assisted by a team of researchers and PhD students that includes Danyal Akarca, Joe Bathelt, Marc Bennett, Giacomo Bignardi, Sarah Bishop, Erica Bottacin, Lara Bridge, Diandra Brkic, Annie Bryant, Sally Butterfield, Elizabeth Byrne, Gemma Crickmore, Edwin Dalmaijer, Fánchea Daly, Tina Emery, Laura Forde, Grace Franckel, Delia Furhmann, Andrew Gadie, Sara Gharooni, Jacalyn Guy, Erin Hawkins, Agnieszka Jaroslawska, Sara Joeghan, Amy Johnson, Jonathan Jones, Silvana Mareva, Elise Ng-Cordell, Sinead O’Brien, Cliodhna O’Leary, Joseph Rennie, Ivan Simpson-Kent, Roma Siugzdaite, Tess Smith, Stephani Uh, Maria Vedechkina, Francesca Woolgar, Natalia Zdorovtsova, Mengya Zhang. The authors wish to thank the many professionals working in children’s services in the South-East and East of England for their support, and to the children and their families for giving up their time to visit the clinic.

## Key Points

• Difficulties with communication, behaviour, and executive function co-occur within and across different neurodevelopmental disorders.
• Our understanding of these co-occurrences, and how they relate to developmental outcomes and neural mechanisms, has arguably been limited by study designs that do not incorporate the heterogeneity within and homogeneity across diagnostic categories.
• We looked at a transdiagnostic cohort of children referred by health and educational professionals and used data-driven community detection to identify three subgroups with distinct profiles of behavioural and communication strengths and difficulties.
• All three data-driven subgroups had more difficulties with academic and socioemotional functioning relative to a demographically matched non-referred group.
• The subgroups could be differentiated from the non-referred sample based on both shared and unique features of neural white matter organisation.
• The three communication and behavioural profiles potentially represent three alternative but related pathways to difficulties with academic and socioemotional functioning.

## Footnotes

1 Subtyping is usually used in the context of applying clustering techniques within a sample of children with the same diagnosis to uncover different presentations of the same disorder (e.g., ADHD-combined and ADHD- inattentive). For clarity, we use the term clustering or subgrouping from this point forward to refer to data-driven clustering approaches applied in diverse samples rather than within a diagnostic category.

