## Supplementary Materials for "Transdiagnostic Profiles of Behaviour and Communication Relate to Academic and Socioemotional Functioning and Neural White Matter Organisation"

### Community detection

#### *Missing data handling*

Missing data was estimated only for the variables used for community detection. The decision was made to keep all other data independent and suitable for external validation. In the referred sample, the missingness across the 24 parent rating subscales used in the community detection analysis ranged from 0.5% to 2.11%. Data was complete for 94% of participants. Missingness levels were similar across boys and girls ($\chi^2$ (1) = 0.06, $p$ = 0.80) and across diagnosed and non-diagnosed participants ($\chi^2$ (1) = 0.0054, $p$ = 0.94). For the non-referred (comparison) sample, 97% of participants had complete data and missingness across variables ranged from 0.63% to 2.53%. Missingness levels were again similar across boys and girls ($\chi^2$ (1) =0.39, $p$ = 0.53).We estimated the missing data separately for each cohort using an identical procedure based on nonparametric missing value imputation using random forests with the *R* package missForest, (version 1.4; Stekhoven & Bühlmann, 2012) implemented with maximum number of iterations set at 10 and number of trees set at 100 (i.e. package defaults). Participant sex and age were also added to the imputation model to improve estimates. The imputed datasets were used in the community detection (referred sample only) and the principal components analyses used to reduce the data (the two separately imputed datasets were combined).

#### *Normality*

Across the 24 subscales used in the community detection, three cases fell outside of ±3.5 SD of the sample mean, and skewness ranged between -1.18 and 1.20. Age was regressed from each subscale using a robust linear model fitting via the R package MASS (Venables & Ripley, 2002). Age-regressed residuals were saved. To limit the influence of univariate outliers, approximate normal distribution percent-ranks ranging from zero to one were estimated for each participant across tasks and the resulting ranks were mapped onto a standard normal distribution using a quantile function (Bignardi et al., 2020; Gregory, 2014). Following this procedure, no cases fell outside the ±3.5 SD of the sample mean range and no variable had skewness values outside the ± 1 range. The normalised residuals were used for all subsequent analyses. As a robustness check the community detection and principal component analyses were also conducted with the non-normalised residuals. No substantial differences regarding the number of subgroups and their characteristics or principal components were noted across the two approaches.

**Figure S1**

*Child-by-child correlation matrix in Fruchterman-Reingold layout color-coded according to the results of the community detection algorithm.*

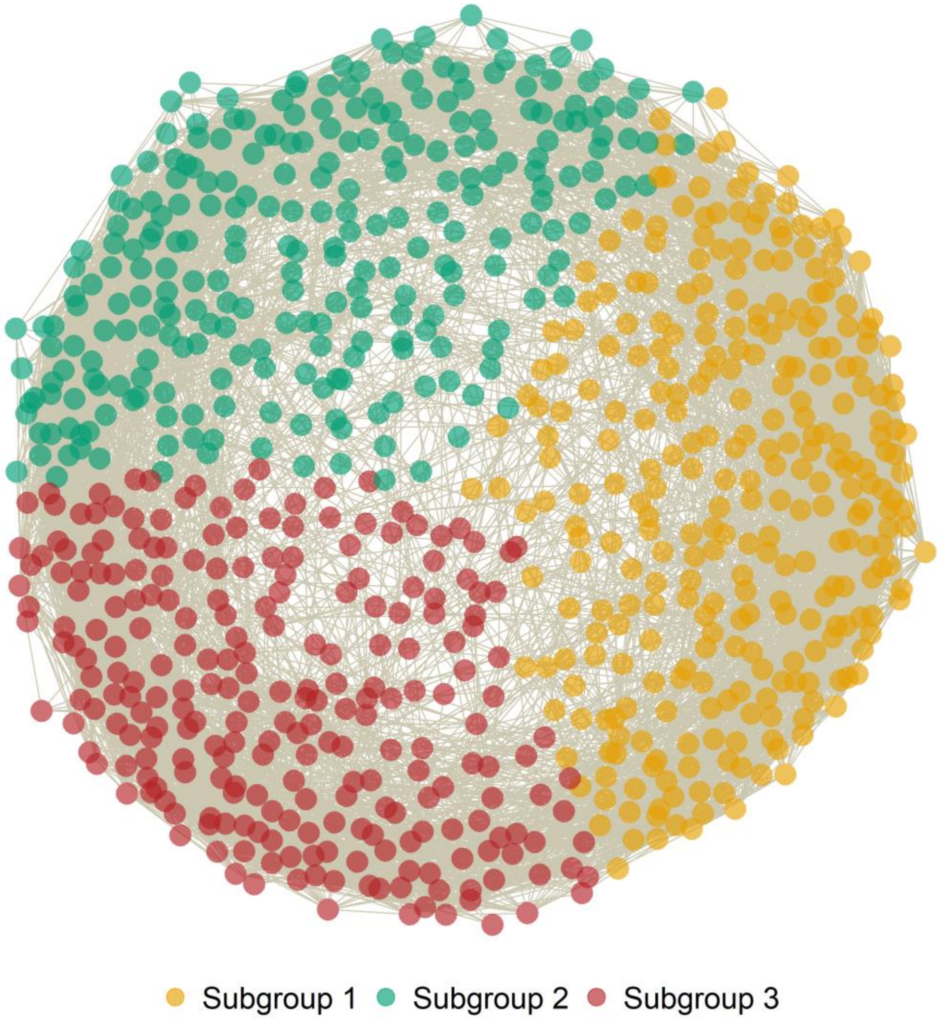


*Note.* Correlation coefficients below the absolute value of .5 were removed for visualisation purposes only. Note that the evaluation of community separation was not guided by the visualisation, but was instead based on the value of the *Q*-statistic, which was derived from the non-thresholded data (i.e., all correlations included).

**Figure S2**

*Pearson correlations across all subscales of Conners. BRIEF, and CCC-2*

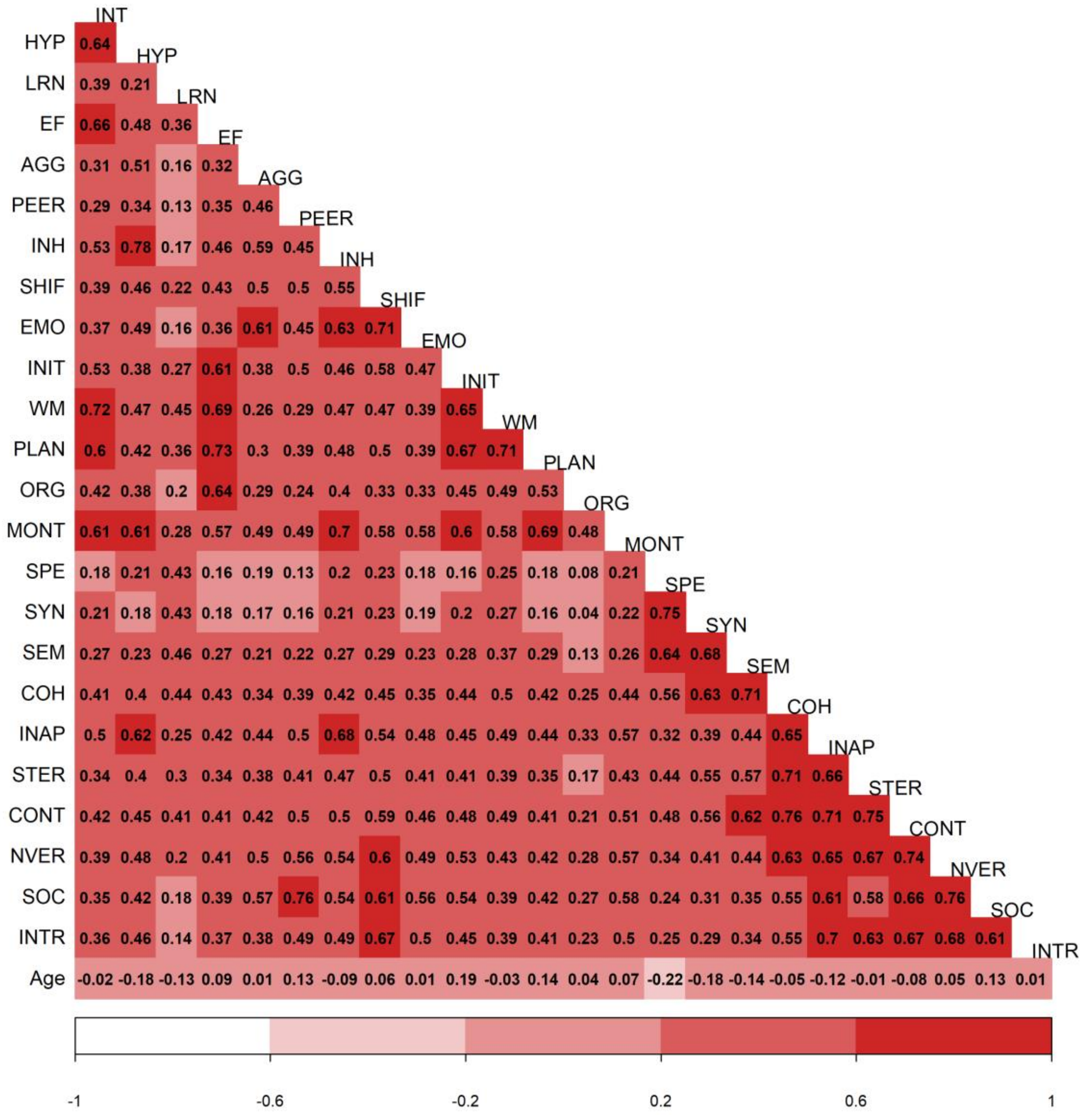


*Note*. Conners (Conners Parent Rating Short Form 3rd Edition) subscales; INT = Inattention; HYP = Hyperactivity/Impulsivity; LRN = Learning Problems; AGG = Aggression; PEER = Peer Relationships; BRIEF (Brief Rating Inventory of Executive Function) subscales: INH = Inhibition; SHIF =Shifting; EMO = Emotional Control; INIT = Initiation; WM = Working memory; PLAN = Planning/Organisation; ORG = Organisation of Materials; MONT = Monitoring; CCC-2 (Children’s Communication Checklist) subscales: SYN = Syntax; SEM = Semantics; COH = Coherence; INAP = Inappropriate Initiation; STER = Stereotyped Language; CONT = Use of Context; NVER= Nonverbal Communication; SOC = Social Relations; INTR = Interests.

**Figure S3**

*Pearson correlations across all subscales of Conners, BRIEF, and CCC-2 after regressing age from each variable*

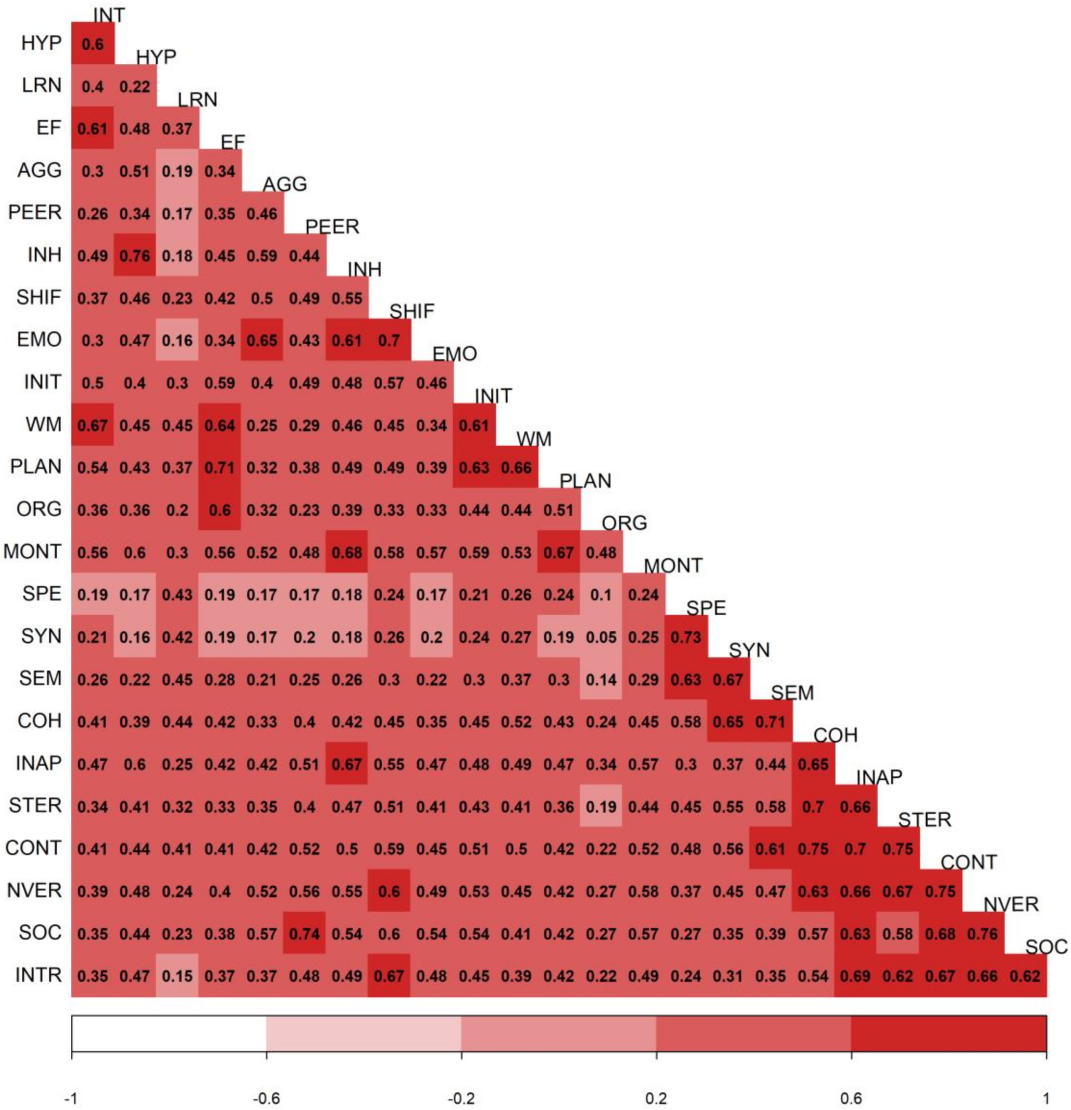


*Note*. Conners (Conners Parent Rating Short Form 3rd Edition) subscales; INT = Inattention; HYP = Hyperactivity/Impulsivity; LRN = Learning Problems; AGG = Aggression; PEER = Peer Relationships; BRIEF (Brief Rating Inventory of Executive Function) subscales: INH = Inhibition; SHIF =Shifting; EMO = Emotional Control; INIT = Initiation; WM = Working memory; PLAN = Planning/Organisation; ORG = Organisation of Materials; MONT = Monitoring; CCC-2 (Children’s Communication Checklist) subscales: SYN = Syntax; SEM = Semantics; COH = Coherence; INAP = Inappropriate Initiation; STER = Stereotyped Language; CONT = Use of Context; NVER= Nonverbal Communication; SOC = Social Relations; INTR = Interests.

**Table S1**

*Descriptive statistics of raw scores derived from the CCC-2, Conners-3, and BRIEF questionnaires and Cohen's d effect size for comparisons across the data-driven subgroups and the comparison sample.*

| | **S1** | | **S2** | | **S3** | | **Nref.** | | **Cohen's *d*** | | | | | |
|---|---|---|---|---|---|---|---|---|---|---|---|---|---|---|
| Scale | *M* | *SD* | *M* | *SD* | *M* | *SD* | *M* | *SD* | S1 vs. S2 | S2 vs. S3 | S1 vs. S3 | S1 vs. Nref. | S2 vs Nref. | S3 vs Nref. |
| INT | 10.5 | 3.9 | 12.7 | 2.7 | 12.0 | 3.0 | 4.1 | 4.0 | 0.6 | -0.2 | 0.4 | **-1.6** | **-2.6** | **-2.3** |
| HYP | 8.9 | 5.6 | 10.7 | 5.4 | 13.6 | 4.5 | 4.4 | 4.5 | 0.3 | 0.6 | 0.9 | -0.9 | **-1.3** | **-2.1** |
| LRN | 10.4 | 3.5 | 10.4 | 3.1 | 8.2 | 3.6 | 2.6 | 3.0 | 0.0 | -0.7 | -0.6 | **-2.4** | **-2.6** | **-1.7** |
| EF | 8.6 | 3.8 | 11.8 | 2.9 | 10.4 | 3.2 | 4.3 | 3.7 | 0.9 | -0.4 | 0.5 | **-1.1** | **-2.3** | **-1.8** |
| AGG | 2.7 | 3.4 | 2.4 | 3.0 | 5.9 | 4.5 | 1.3 | 2.3 | -0.1 | 0.9 | 0.8 | -0.4 | -0.4 | **-1.2** |
| PEER | 5.1 | 4.2 | 3.8 | 3.8 | 8.0 | 4.2 | 1.6 | 2.9 | -0.3 | **1.1** | 0.7 | **-0.9** | -0.6 | **-1.7** |
| INH | 19.3 | 5.9 | 20.5 | 6.0 | 25.6 | 4.5 | 14.4 | 4.8 | 0.2 | **1.0** | **1.2** | **-0.9** | **-1.1** | **-2.4** |
| SHIF | 16.3 | 4.3 | 16.0 | 4.0 | 19.2 | 3.6 | 11.9 | 3.7 | -0.1 | 0.8 | 0.7 | **-1.1** | **-1.1** | **-2.0** |
| EMO | 20.6 | 5.4 | 20.8 | 5.5 | 25.5 | 4.3 | 16.1 | 4.7 | 0.0 | **1.0** | **1.0** | -0.9 | -0.9 | **-2.1** |
| INIT | 17.0 | 3.6 | 18.0 | 3.1 | 18.3 | 3.0 | 12.8 | 3.5 | 0.3 | 0.1 | 0.4 | **-1.2** | **-1.6** | **-1.7** |
| WM | 24.6 | 4.5 | 27.0 | 3.3 | 25.7 | 3.8 | 15.6 | 5.1 | 0.6 | -0.4 | 0.3 | **-1.9** | **-2.8** | **-2.3** |
| PLAN | 27.0 | 5.4 | 30.6 | 4.2 | 29.6 | 4.4 | 19.3 | 6.0 | 0.8 | -0.2 | 0.5 | **-1.4** | **-2.3** | **-2.0** |
| ORG | 13.1 | 3.5 | 16.0 | 2.4 | 15.1 | 2.8 | 12.0 | 3.3 | **1.0** | -0.4 | 0.6 | -0.3 | **-1.5** | **-1.0** |
| MONT | 17.7 | 3.9 | 19.3 | 3.0 | 20.4 | 2.9 | 13.0 | 3.8 | 0.5 | 0.4 | 0.8 | **-1.2** | **-1.9** | **-2.3** |
| SPE | 7.8 | 5.4 | 3.7 | 4.1 | 3.5 | 3.9 | 1.6 | 2.7 | -0.8 | -0.1 | -0.9 | **-1.3** | -0.6 | -0.5 |
| SYN | 8.1 | 5.1 | 3.3 | 3.5 | 3.5 | 3.2 | 1.2 | 2.3 | **-1.1** | 0.1 | **-1.1** | **-1.6** | -0.7 | -0.8 |
| SEM | 10.6 | 4.7 | 6.6 | 4.4 | 6.7 | 3.9 | 1.9 | 2.5 | -0.9 | 0.0 | -0.9 | **-2.2** | **-1.2** | **-1.4** |
| COH | 10.3 | 5.1 | 6.6 | 4.7 | 8.6 | 4.7 | 2.2 | 3.0 | -0.8 | 0.4 | -0.4 | **-1.8** | **-1.0** | **-1.5** |
| INAP | 10.6 | 5.6 | 8.6 | 5.3 | 13.5 | 5.1 | 3.7 | 3.4 | -0.4 | **1.0** | 0.5 | **-1.4** | **-1.0** | **-2.2** |
| STER | 7.2 | 4.6 | 4.1 | 3.4 | 6.9 | 4.0 | 1.9 | 2.5 | -0.8 | 0.8 | -0.1 | **-1.3** | -0.7 | **-1.4** |
| CONT | 11.0 | 5.3 | 6.9 | 4.9 | 10.9 | 5.2 | 2.7 | 3.3 | -0.8 | 0.8 | 0.0 | **-1.8** | -0.9 | **-1.8** |
| NVER | 8.1 | 5.0 | 5.5 | 4.4 | 9.9 | 4.8 | 2.5 | 3.3 | -0.6 | 0.9 | 0.4 | **-1.2** | -0.7 | **-1.7** |
| SOC | 6.6 | 5.0 | 4.4 | 4.0 | 9.2 | 4.5 | 2.0 | 2.9 | -0.5 | **1.1** | 0.6 | **-1.1** | -0.7 | **-1.8** |
| INTR | 9.2 | 4.5 | 7.1 | 3.9 | 11.5 | 4.2 | 4.7 | 3.3 | -0.5 | **1.1** | 0.5 | **-1.1** | -0.7 | **-1.8** |

*Note.* Conners (Conners Parent Rating Short Form 3rd Edition) subscales; INT = Inattention; HYP = Hyperactivity/Impulsivity; LRN = Learning Problems; AGG = Aggression; PEER = Peer Relationships; BRIEF (Brief Rating Inventory of Executive Function) subscales: INH = Inhibition; SHIF =Shifting; EMO = Emotional Control; INIT = Initiation; WM = Working memory; PLAN = Planning/Organisation; ORG = Organisation of Materials; MONT = Monitoring; CCC-2 (Children's Communication Checklist) subscales: SYN = Syntax; SEM = Semantics; COH = Coherence; INAP = Inappropriate Initiation; STER = Stereotyped Language; CONT = Use of Context; NVER= Nonverbal Communication; SOC = Social Relations; INTR = Interests. S1 = Subgroup 1, S2 = Subgroup 2, S3 = Subgroup 3, Nref. = Non-referred group. Cohen's $d$ > |1| presented in bold.

### Data reduction: Principal component analyses

The parent rating data from the CCC-2, Conners, and BRIEF ratings for each cohort was combined. Principal component analyses (PCA) was then used separately on the communication (i.e. all CCC-2 ratings) and behavioural data (i.e. Conners and BRIEF ratings). We used an orthogonal rotation (i.e. varimax) to maximise differences between the extracted components. The maximum number of components to retain was chosen based on the results of parallel analysis. All component structures from one to the maximum suggested in the parallel analysis were considered. Component structures were considered interpretable if each component had at least three primary loadings (highest loading ≥0.35 and at least 0.10 greater than all other loadings). For both the behavioural and communication data, the two-component solution was the one that met these criteria. For the CCC-2 data, the two principal components in combination explained 80% of the variance. The first component captured pragmatic communication (accounting for 46% of variance), and the second structural language skills (accounting for 34% of variance). In the behavioural data, the two components cumulatively explained 68% of the variance. The first component (accounting for 37% of variance) was labelled cool executive function (EF), and the second component (accounting for 31% of variance) hot EF. The components resembled the clusters of difficulties identified by Mareva & Holmes (2019). The results of the parallel analyses are presented in Figure S4 and Figure S5 and the loadings from each PCA are presented in Figure 1b and Figure 1c in the manuscript. Component scores were found using the regression method (for details see Revelle, 2019) and were saved for subsequent analyses. Note that repeating the PCA separately for the referred and non-referred samples supported the same conclusions.

As a sensitivity check, we also repeated the PCA on the combined communication and behaviour data (CCC-2, Conners and BRIEF). In this case, parallel analyses suggested that three components could be extracted (Figure S6). The components were extracted using a varimax rotation and collectively explained 71% of the variance. The loadings are presented in Figure S7. In this case, a single factor captured both pragmatics and hot EFs, a second factor captured cool EF, and the third factor captured structural language. The means and standard deviations of the component scores derived from this analysis are presented in Table S2 and support the same conclusions as those based on the separate PCA models reported in the manuscript.

**Figure S4**

*Parallel analysis of the CCC-2 subscales*

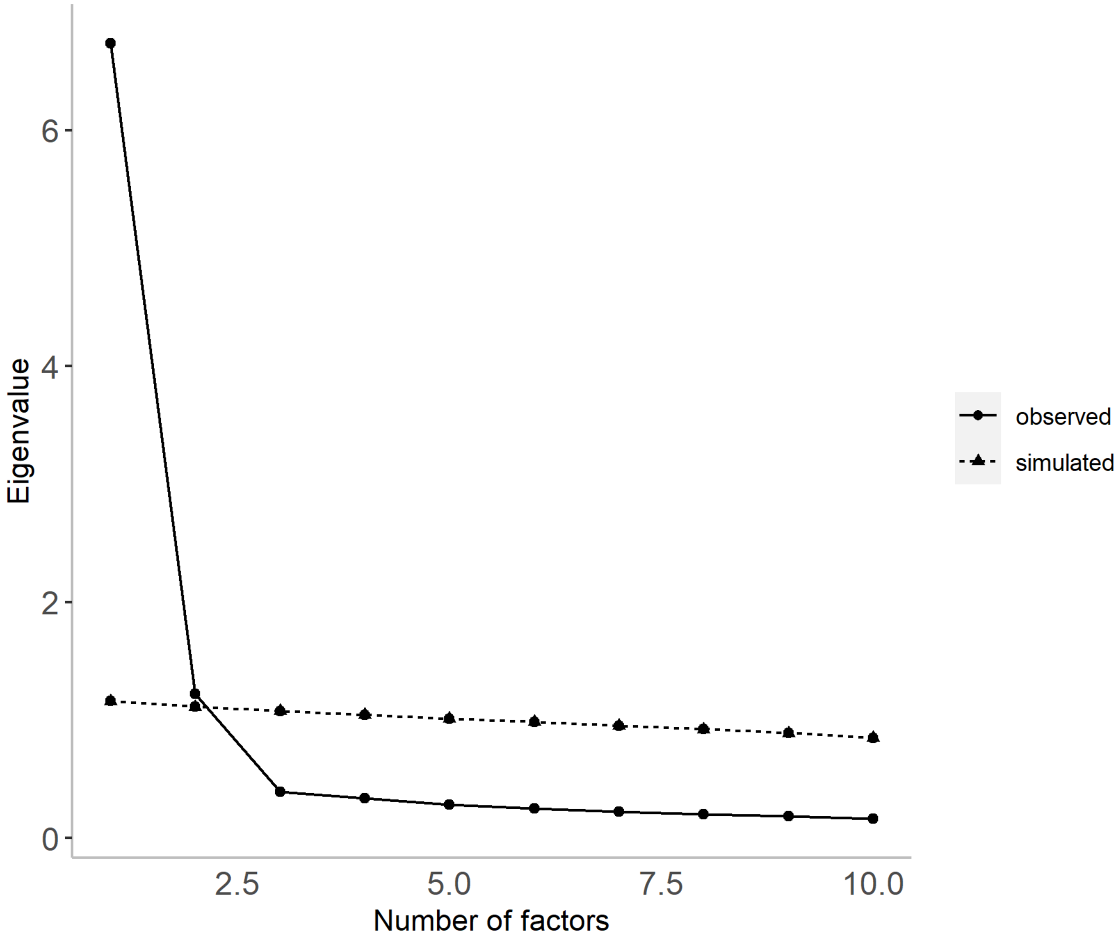


*Note.* The simulated line is the top of 95% confidence interval around simulated eigenvalues. CCC-2 = Children communications checklist.

**Figure S5**

*Parallel analysis of the Conners and BRIEF subscales*

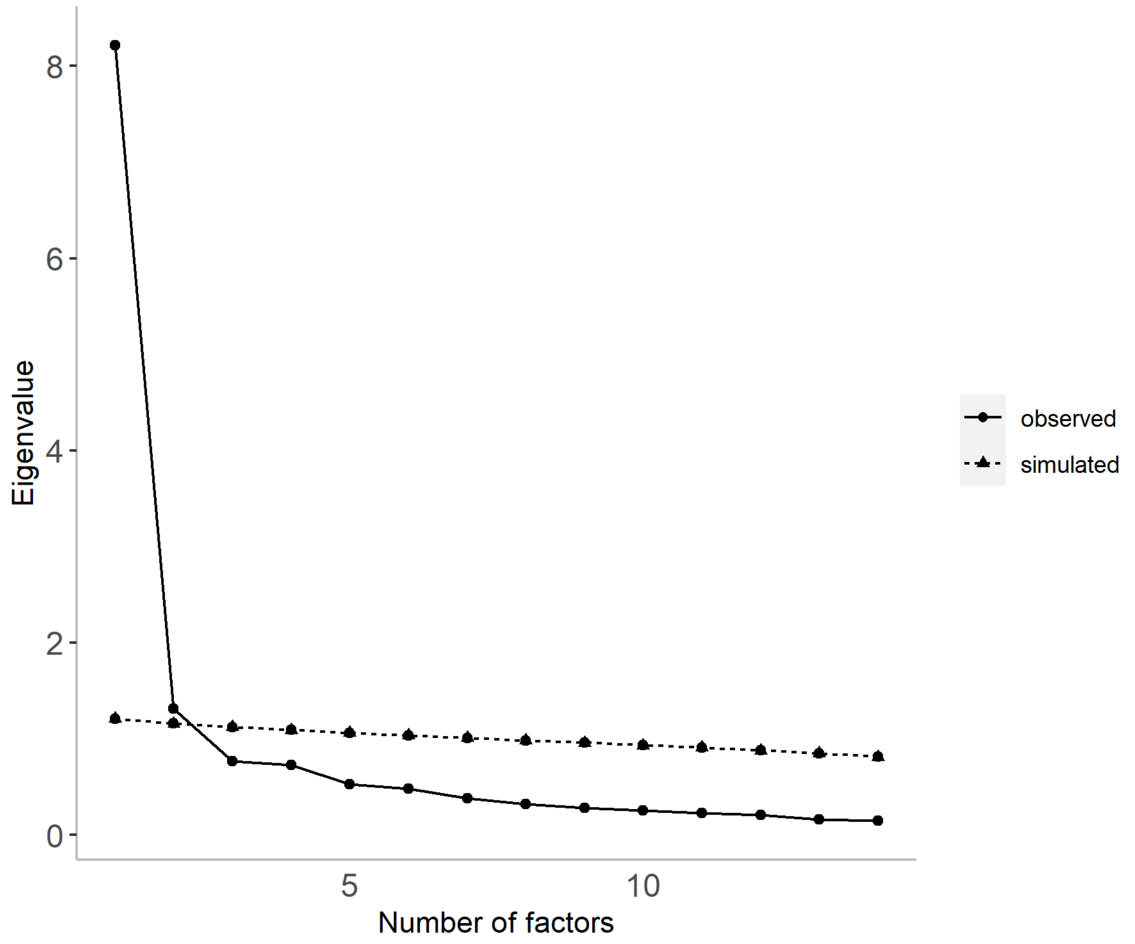


*Note.* The simulated line is the top of 95% confidence interval around simulated eigenvalues. Conners = Conners Parent Rating Short Form 3rd Edition. BRIEF = Brief Rating Inventory of Executive Function.

**Figure S6**

*Parallel analysis of the CCC-2, Conners and BRIEF subscales*

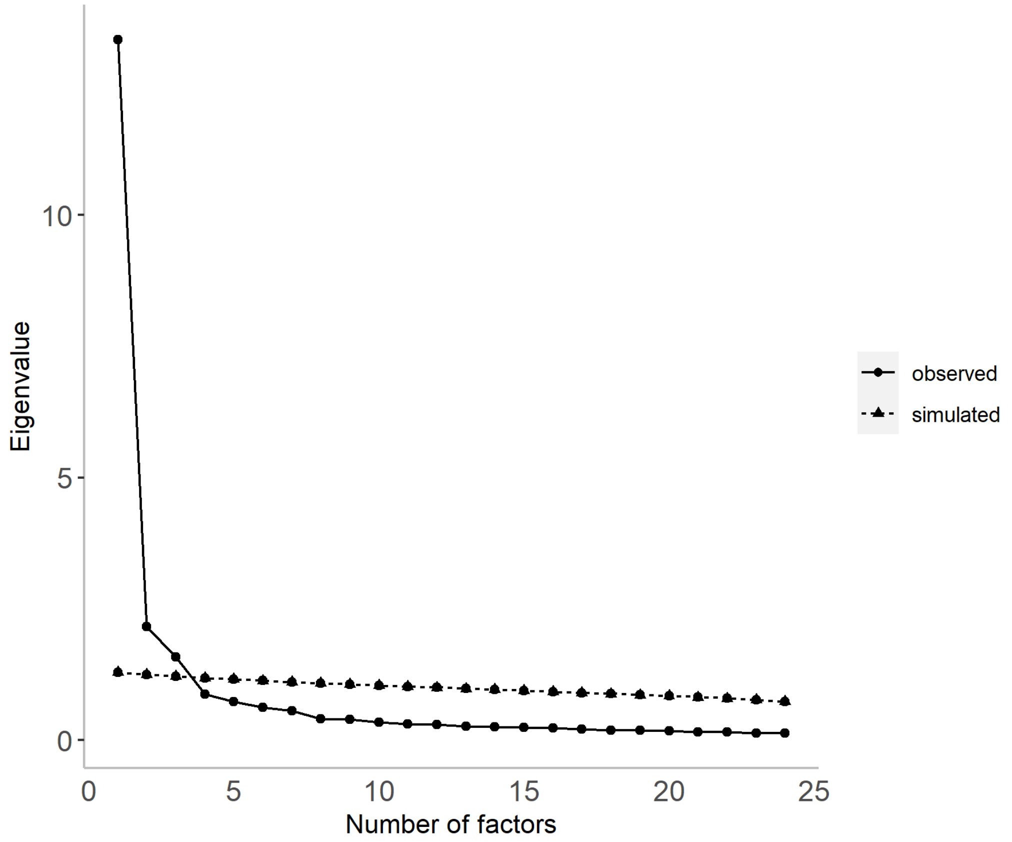


*Note.* The simulated line is the top of 95% confidence interval around simulated eigenvalues. Conners = Conners Parent Rating Short Form 3rd Edition. BRIEF = Brief Rating Inventory of Executive Function. CCC-2 = Children communications checklist.

**Figure S7**

*Varimax loadings from Principal component analyses (PCA) of the Conners, CCC-2, and BRIEF subscales based on combined data from the referred and comparison samples.*

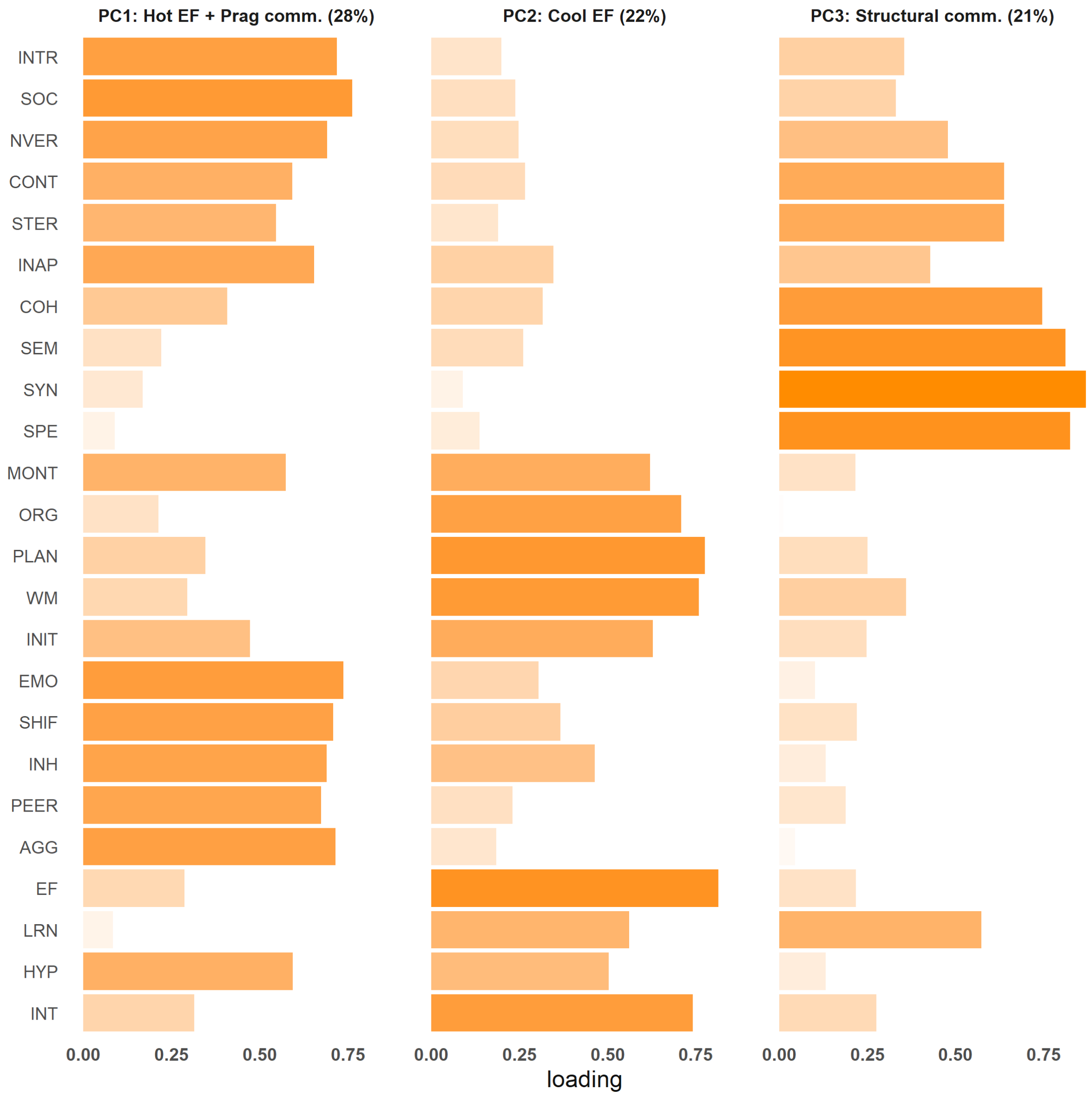


*Note*. Conners = Conners Parent Rating Short Form 3rd Edition. BRIEF = Brief Rating Inventory of Executive Function. CCC-2 = Children communications checklist.

**Table S2**

*Descriptive statistics across the data-driven subgroups and the comparison sample for all behavioural measures and principal components*

| Measure | Subgroup 1 | | | Subgroup 2 | | | Subgroup 3 | | | Non-referred | | |
|---|---|---|---|---|---|---|---|---|---|---|---|---|
| | *N* | *M* | *SD* | *N* | *M* | *SD* | *N* | *M* | *SD* | *N* | *M* | *SD* |
| SDQ: Emotion problems (raw) | 297 | 4.22 | 2.70 | 259 | 3.96 | 2.76 | 240 | 5.17 | 2.76 | 157 | 1.86 | 2.27 |
| SDQ: Conduct problems (raw) | 297 | 2.84 | 2.29 | 259 | 2.95 | 2.34 | 241 | 4.93 | 2.46 | 157 | 1.47 | 1.75 |
| SDQ: Peer problems (raw) | 297 | 3.27 | 2.59 | 259 | 2.40 | 2.30 | 241 | 4.74 | 2.42 | 157 | 1.24 | 1.81 |
| SDQ: Prosocial (raw) | 297 | 7.14 | 2.41 | 259 | 7.42 | 2.20 | 241 | 5.76 | 2.17 | 157 | 8.15 | 1.84 |
| SDQ: Emotion problems (res.) | 297 | 0.22 | 2.68 | 259 | -0.05 | 2.75 | 240 | 1.2 | 2.74 | 157 | -2.17 | 2.28 |
| SDQ: Conduct problems (res.) | 297 | -0.33 | 2.29 | 259 | -0.23 | 2.34 | 241 | 1.75 | 2.46 | 157 | -1.7 | 1.75 |
| SDQ: Peer problems (res.) | 297 | 0.2 | 2.53 | 259 | -0.69 | 2.27 | 241 | 1.72 | 2.37 | 157 | -1.89 | 1.84 |
| SDQ: Prosocial (res.) | 297 | 0.11 | 2.4 | 259 | 0.39 | 2.19 | 241 | -1.29 | 2.17 | 157 | 1.13 | 1.84 |
| WIAT-II: Numerical operations | 265 | 81.25 | 15.74 | 233 | 85.38 | 15.02 | 227 | 90.58 | 17.91 | 157 | 115.52 | 18.11 |
| WIAT-II: Word reading | 294 | 80.75 | 15.64 | 255 | 88.49 | 16.07 | 236 | 93.17 | 16.62 | 156 | 108.68 | 12.83 |
| WASI-II: Matrix reasoning | 300 | 40.96 | 8.77 | 261 | 43.48 | 9.33 | 242 | 45.14 | 10.26 | 158 | 53.28 | 9.14 |
| PC: Structural language (CCC-2) | 300 | -0.82 | 0.78 | 261 | 0.08 | 0.85 | 244 | 0.47 | 0.79 | 158 | 0.7 | 0.82 |
| PC: Pragmatic Communication (CCC-2) | 300 | 0.01 | 0.9 | 261 | 0.24 | 0.79 | 244 | -0.83 | 0.78 | 158 | 0.87 | 0.82 |
| PC: Cool EF (Conners + BRIEF) | 300 | -0.02 | 0.8 | 261 | -0.8 | 0.78 | 244 | 0.12 | 0.72 | 158 | 1.19 | 0.76 |
| PC: Hot EF (Conners + BRIEF) | 300 | 0.16 | 0.89 | 261 | 0.41 | 0.87 | 244 | -0.95 | 0.74 | 158 | 0.48 | 0.75 |
| PC: Hot EF + Pragmatic Communication (CCC-2, Conners, BRIEF) | 300 | 0.13 | 0.88 | 261 | 0.53 | 0.81 | 244 | -1.02 | 0.67 | 158 | 0.44 | 0.74 |
| PC: Cool EF (CCC-2, Conners, BRIEF) | 300 | 0.36 | 0.72 | 261 | -1 | 0.69 | 244 | -0.03 | 0.67 | 158 | 1.02 | 0.86 |
| PC: Structural language (CCC-2, Conners, BRIEF) | 300 | -0.94 | 0.72 | 261 | 0.26 | 0.80 | 244 | 0.38 | 0.72 | 158 | 0.77 | 0.83 |

*Note.* WIAT-II = Wechsler Individual Achievement Test-II; WASI-II = Wechsler Abbreviated Scale of Intelligence II; SDQ = Strengths and difficulties questionnaire, parent-report. res. = age-regressed residual. Conners = Conners Parent Rating Short Form 3$^{rd}$ Edition. BRIEF = Brief Rating Inventory of Executive Function. CCC-2 = Children communications checklist. PC = Principal Component. For all principal components, the text in parentheses indicates from which measures the component was identified.

### Magnetic resonance imaging pre-processing

*Anatomical data pre-processing*

The T1-weighted (T1w) image was corrected for intensity non-uniformity (INU) using N4BiasFieldCorrection (Tustison et al., 2010) and used as T1w-reference throughout the workflow. The T1w-reference was then skull-stripped using antsBrainExtraction.sh (ANTs 2.3.1), using OASIS as target template. Spatial normalization to the ICBM 152 Nonlinear Asymmetrical template version 2009c (RRID:SCR_008796, Fonov et al., 2009) was performed through nonlinear registration with antsRegistration (ANTs 2.3.1, RRID:SCR_004757, Avants et al., 2008), using brain-extracted versions of both T1w volume and template. Brain tissue segmentation of cerebrospinal fluid, white-matter, and grey-matte was performed on the brain-extracted T1w using FAST (FSL 6.0.3:b862cdd5, RRID:SCR_002823, Zhang et al., 2001).

*Diffusion data pre-processing*

Any images with a b-value less than 100 s/mm^2 were treated as a *b*=0 image. MP-PCA denoising as implemented in MRtrix3's *dwidenoise* (Veraart et al., 2016) was applied with a 5-voxel window. After MP-PCA, B1 field inhomogeneity was corrected using *dwibiascorrect* from MRtrix3 with the N4 algorithm (Tustison et al., 2010). After B1 bias correction, the mean intensity of the DWI series was adjusted so all the mean intensity of the b=0 images matched across each separate DWI scanning sequence. FSL (version 6.0.3:b862cdd5)'s eddy was used for head motion correction and Eddy current correction (Andersson & Sotiropoulos, 2016). Eddy was configured with a *q*-space smoothing factor of 10, a total of 5 iterations, and 1000 voxels used to estimate hyperparameters. A linear first level model and a linear second level model were used to characterise Eddy current-related spatial distortion. *Q*-space coordinates were forcefully assigned to shells. Field offset was attempted to be separated from subject movement. Shells were aligned post-eddy. Eddy's outlier replacement was run (Andersson et al., 2016). Data were grouped by slice, only including values from slices determined to contain at least 250 intracerebral voxels. Groups deviating by more than 4 standard deviations from the prediction had their data replaced with imputed values. Final interpolation was performed using the "jac" method (Jenkinson et al., 2012). Framewise displacement was calculated using the implementation *Nipype* following the definitions by Power and colleagues (2014). Many internal operations of the QSIPrep pipeline use *Nilearn* 0.7.0 (Abraham et al., 2014) and *Dipy* (Garyfallidis et al., 2014). More details are available online at: https://qsiprep.readthedocs.io .

*DSI Studio Reconstruction*

Diffusion orientation distribution functions were reconstructed using generalised q-sampling imaging (GQI, Yeh et al., 2010) with a ratio of mean diffusion distance of 1.25. We used the standard GQI plus deterministic tractography pipeline (*dsi_studio_gqi*) (Yeh et al., 2013). 5 million streamlines were created with a maximum length of 250mm, minimum length of 30mm, random seeding with a step size of 1mm.

*Network-Based Statistics*

To investigate differences in the connections between regions we used network-based statistics, which in many cases offers greater statistical power compared to traditional correction methods (Zalesky et al., 2010). This involved comparing each data-driven subgroup to the comparison group in a series of between-group *t*-tests for any pair of regions. For each group comparison, a pre-determined threshold of Cohen's *d* effect size ≥ 0.4 (i.e., $t = 2.8$) was used to select which pairs of regions would be chosen for further analyses. Pairs of regions that met this condition were then used to identify subnetworks for which a path could be found between any two regions. A family-wise error (FWE)-corrected *p*-value was then obtained for each subnetwork via permutation tests ($N = 1000$). For each permutation, the children were randomly exchanged between the data-driven subgroup and the comparison group. The size of the subnetwork can be measured in terms of its extent (i.e., the total number of connections) or intensity (i.e., sum of test statistic values across all connections). The probability of observing a subnetwork of a given extent or intensity is estimated as the proportion of permutations for which the largest subnetwork was the same size or greater. In the current analyses, we focused on intensity and considered subnetworks which met the FWE-corrected threshold of $p < 0.01$ significantly different between groups. Notably, the FWE-correction does not allow inferences to be drawn at the level of individual connections. Age, gender, and average frame displacement were used as covariates in all analyses to mitigate their potential confounding effects. To ensure the robustness of the results, we repeated all procedures by varying the target *t*-threshold, exploring the extent-based measure of size, and setting a more liberal FWE-correction ($p < .05$).

**Figure S8**

*Comparison of the global organisation of the white matter connectome between the data-driven subgroups and the comparison sample.*

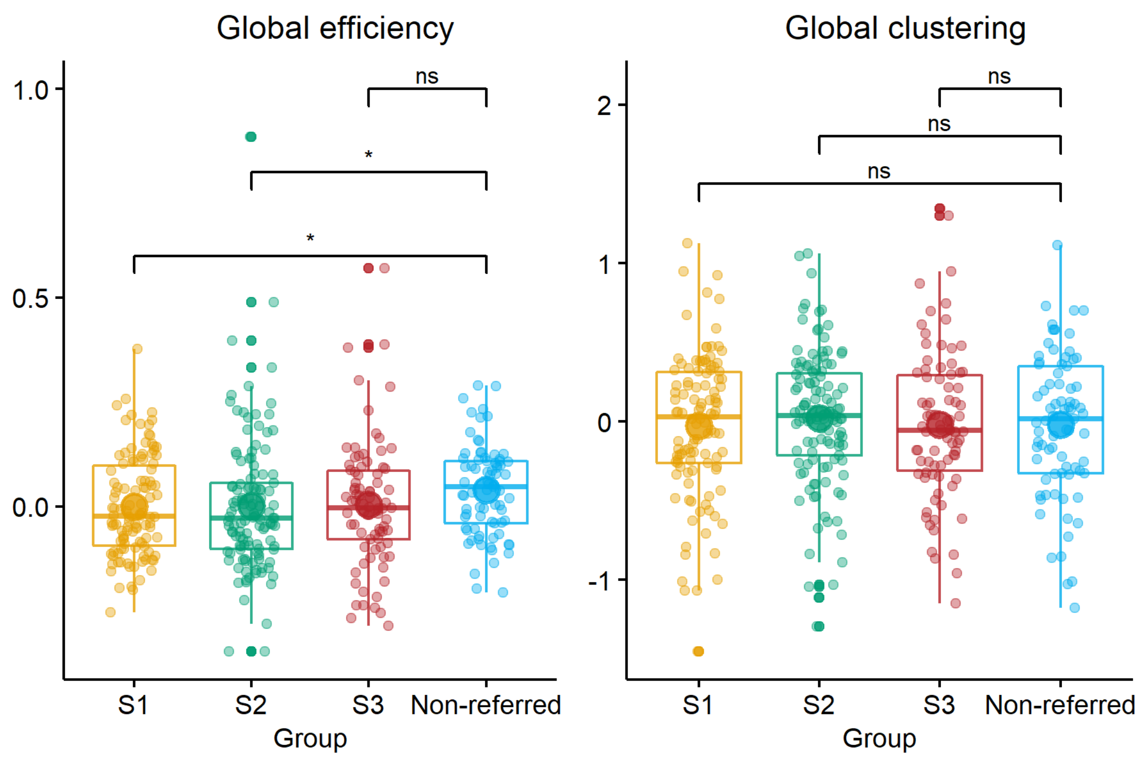


*Note.* Comparisons are based on 10% trimmed means to account for the presence of outliers and *p*-values are adjusted with false discovery rate correction. S1 = Subgroup 1 (most severe structural language difficulties); S2 = Subgroup 2 (most severe *cool* executive difficulties); S3 = Subgroup 3 (most severe difficulties with *hot* executive skills & pragmatic communication).

ns = not significant.

* $p_{corrected} < .05$

**Figure S9**

*For each subgroup the overall network connectivity strength within the intrinsic connectivity networks (defined by Yeo et al., 2011) and the subcortex was compared to connectivity observed in the comparison sample. The location of the dots shows the estimated effect size based on a 10% trimmed mean comparison used to account for the presence of outliers, the colour of the dots denotes whether the corresponding false-discovery-rate adjusted p-value was significant. The width of the lines corresponds to the 95% Confidence intervals.*

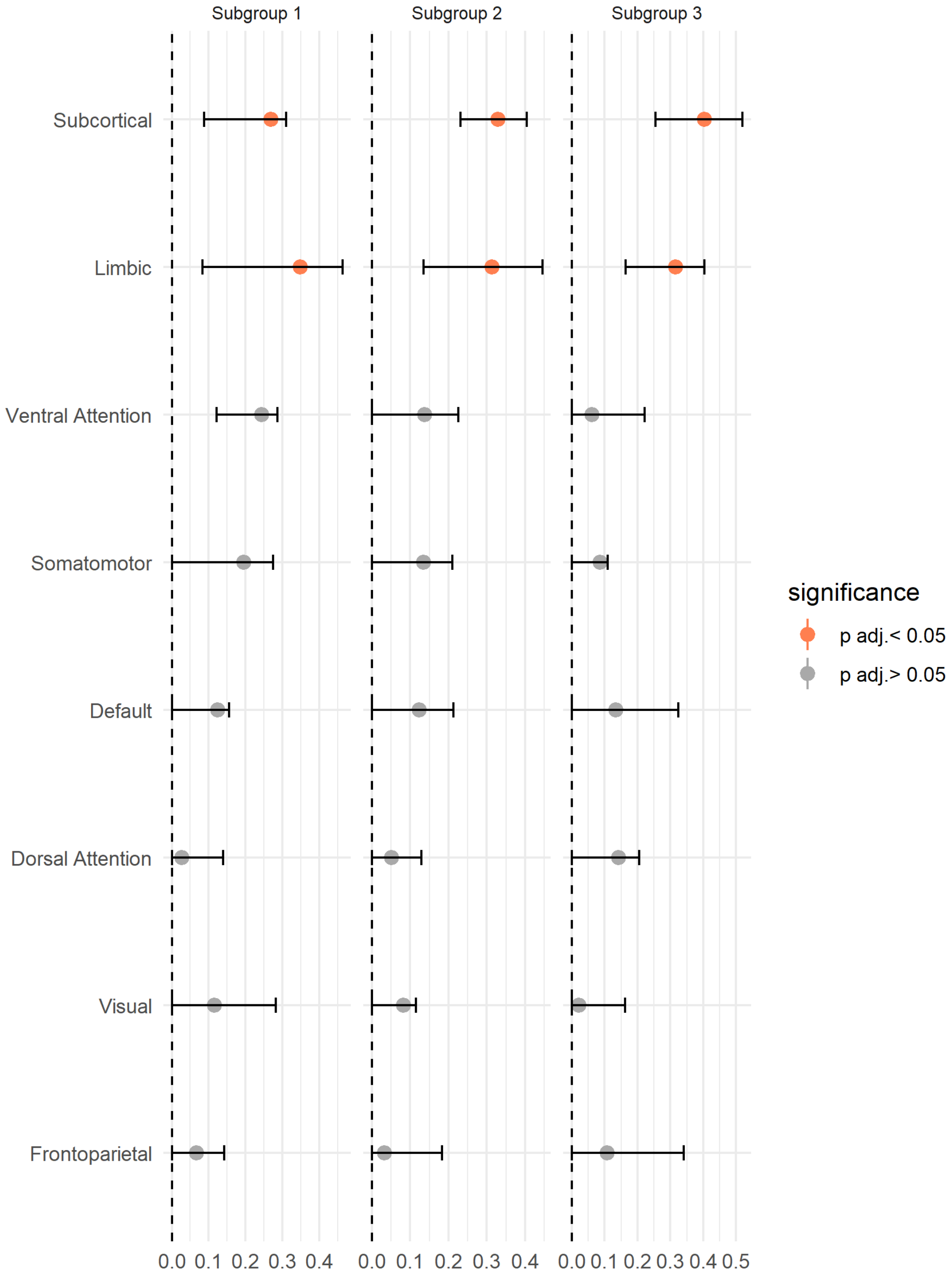


*Note.* Subgroup 1: most severe structural language difficulties; Subgroup 2: most severe cool executive difficulties; Subgroup 3: most severe difficulties with hot executive skills & pragmatic communication.

**Table S3**

*Subcortical and limbic network sub-regions that showed a significant reduction in connection strength relative to the comparison sample in only one of the data-driven subgroups. Means represent age, gender, and motion-corrected residuals and are 10% trimmed to account for outliers.*

| Sub-region | $M_{trim}$ | Non-referred $M_{trim}$ | $p_{adj.}$ | Effect size | Low 95% CI | Up 95% CI | Gyrus | Hemisphere |
|---|---|---|---|---|---|---|---|---|
| | **Subgroup 1** | | | | | | | |
| ventromedial putamen | -1.77 | 2.12 | 0.03 | 0.27 | 0.17 | 0.30 | Basal Ganglia | Left |
| intermediate lateral area 20 | -3.28 | -1.64 | 0.04 | 0.24 | 0.12 | 0.32 | Inferior Temporal Gyrus | Right |
| | **Subgroup 2** | | | | | | | |
| lateral amygdala | -1.48 | 0.00 | 0.02 | 0.27 | 0.19 | 0.38 | Amygdala | Left |
| caudoventral of area 20 | -4.59 | 2.62 | 0.004 | 0.36 | 0.18 | 0.42 | Inferior Temporal Gyrus | Right |
| dorsal caudate | 1.12 | -3.25 | 0.04 | 0.23 | 0.15 | 0.30 | Basal Ganglia | Left |
| area TI (temporal agranular insular cortex) | -1.79 | 1.82 | 0.04 | 0.21 | 0.08 | 0.36 | Parahippocampal Gyrus | Right |
| medial area 38 | -5.63 | 3.92 | 0.04 | 0.22 | 0.13 | 0.32 | Superior Temporal Gyrus | Right |
| | **Subgroup 3** | | | | | | | |
| dorsolateral putamen | -6.42 | 1.29 | 0.03 | 0.26 | 0.10 | 0.35 | Basal Ganglia | Left |
| rostroventral area 24 | -1.53 | 1.03 | 0.02 | 0.27 | 0.15 | 0.36 | Cingulate Gyrus | Left |
| caudal temporal thalamus | -1.25 | 0.54 | 0.01 | 0.30 | 0.12 | 0.39 | Thalamus | Right |

*Note.* Subgroup 1: most severe structural language difficulties; Subgroup 2: most severe cool executive difficulties; Subgroup 3: most severe difficulties with hot executive skills & pragmatic communication.

**Table S4**

*Subcortical and limbic network sub-regions that showed a significant reduction in connection strength relative to the comparison sample in multiple data-driven subgroups. Means represent age, gender, and motion-corrected residuals and are 10% trimmed to account for outliers.*

| Sub-region | $M_{trim}$ | Subgroup | Nref. $M_{trim}$ | p. | Effect size | Low 95% CI | Up 95% | Gyrus | Hemi. |
|---|---|---|---|---|---|---|---|---|---|
| caudal hippocampus | -0.77 | 1 | 10.31 | 0.03 | 0.26 | 0.19 | 0.42 | Hippocampus | L |
| caudal hippocampus | -5.61 | 2 | 10.31 | 0.00 | 0.42 | 0.27 | 0.49 | Hippocampus | L |
| caudal hippocampus | -5.97 | 3 | 10.31 | 0.01 | 0.33 | 0.14 | 0.52 | Hippocampus | L |
| caudoventral of area 20 | -2.64 | 1 | 3.75 | 0.03 | 0.28 | 0.09 | 0.43 | Inferior Temporal Gyrus | L |
| caudoventral of area 20 | -4.59 | 2 | 2.62 | 0.00 | 0.36 | 0.18 | 0.42 | Inferior Temporal Gyrus | R |
| caudoventral of area 20 | -0.76 | 3 | 3.75 | 0.03 | 0.23 | 0.12 | 0.39 | Inferior Temporal Gyrus | L |
| globus pallidus | 0.03 | 1 | 3.15 | 0.04 | 0.28 | 0.19 | 0.37 | Basal Ganglia | L |
| globus pallidus | -1.47 | 2 | 3.15 | 0.02 | 0.30 | 0.14 | 0.46 | Basal Ganglia | L |
| globus pallidus | -4.85 | 2 | 4.00 | 0.02 | 0.28 | 0.11 | 0.30 | Basal Ganglia | R |
| globus pallidus | -4.34 | 3 | 3.15 | 0.00 | 0.47 | 0.28 | 0.63 | Basal Ganglia | L |
| globus pallidus | -6.10 | 3 | 4.00 | 0.01 | 0.31 | 0.10 | 0.45 | Basal Ganglia | R |
| intermediate ventral area 20 | -1.05 | 1 | 1.55 | 0.04 | 0.22 | 0.13 | 0.28 | Inferior Temporal Gyrus | L |
| intermediate ventral area 20 | -1.46 | 2 | 1.55 | 0.04 | 0.21 | 0.04 | 0.35 | Inferior Temporal Gyrus | L |
| intermediate ventral area 20 | -1.22 | 3 | 1.55 | 0.02 | 0.27 | 0.15 | 0.39 | Inferior Temporal Gyrus | L |
| medial area 11 | -4.25 | 2 | 4.97 | 0.04 | 0.24 | 0.11 | 0.31 | Orbital Gyrus | L |
| medial area 11 | -6.02 | 3 | 4.97 | 0.01 | 0.34 | 0.20 | 0.57 | Orbital Gyrus | L |
| medial pre-frontal thalamus | -1.72 | 1 | 2.94 | 0.04 | 0.22 | 0.08 | 0.38 | Thalamus | L |
| medial pre-frontal thalamus | -0.87 | 2 | 2.94 | 0.04 | 0.23 | 0.13 | 0.36 | Thalamus | L |
| medial pre-frontal thalamus | -2.39 | 3 | 2.94 | 0.03 | 0.26 | 0.19 | 0.44 | Thalamus | L |
| nucleus accumbens | 1.23 | 1 | 4.93 | 0.04 | 0.26 | 0.16 | 0.35 | Basal Ganglia | L |
| nucleus accumbens | -2.18 | 2 | 4.93 | 0.01 | 0.32 | 0.22 | 0.39 | Basal Ganglia | L |
| nucleus accumbens | -5.52 | 3 | 4.93 | 0.01 | 0.32 | 0.21 | 0.47 | Basal Ganglia | L |
| rostroventral area 20 | -1.73 | 1 | 9.17 | 0.04 | 0.22 | 0.10 | 0.34 | Fusiform Gyrus | L |
| rostroventral area 20 | -7.99 | 2 | 9.17 | 0.01 | 0.34 | 0.14 | 0.49 | Fusiform Gyrus | L |
| rostroventral area 20 | -4.33 | 3 | 9.17 | 0.01 | 0.31 | 0.15 | 0.47 | Fusiform Gyrus | L |
| ventral caudate | -4.80 | 1 | 5.61 | 0.03 | 0.32 | 0.29 | 0.42 | Basal Ganglia | L |
| ventral caudate | -2.64 | 1 | 2.23 | 0.04 | 0.24 | 0.06 | 0.26 | Basal Ganglia | R |
| ventral caudate | -1.19 | 2 | 5.61 | 0.04 | 0.20 | 0.03 | 0.26 | Basal Ganglia | L |
| ventral caudate | -2.32 | 2 | 2.23 | 0.04 | 0.21 | 0.09 | 0.34 | Basal Ganglia | R |
| ventral caudate | -3.85 | 3 | 5.61 | 0.01 | 0.39 | 0.20 | 0.50 | Basal Ganglia | L |

*Note.* Subgroup 1: most severe structural language difficulties; Subgroup 2: most severe cool executive difficulties; Subgroup 3: most severe difficulties with hot executive skills & pragmatic communication. Nref. = non-referred.

**Figure S10**

*Subnetworks identified as significantly weaker in Subgroup 3 relative to the comparison sample when a more liberal family-wise error correction (p<.05) was applied*

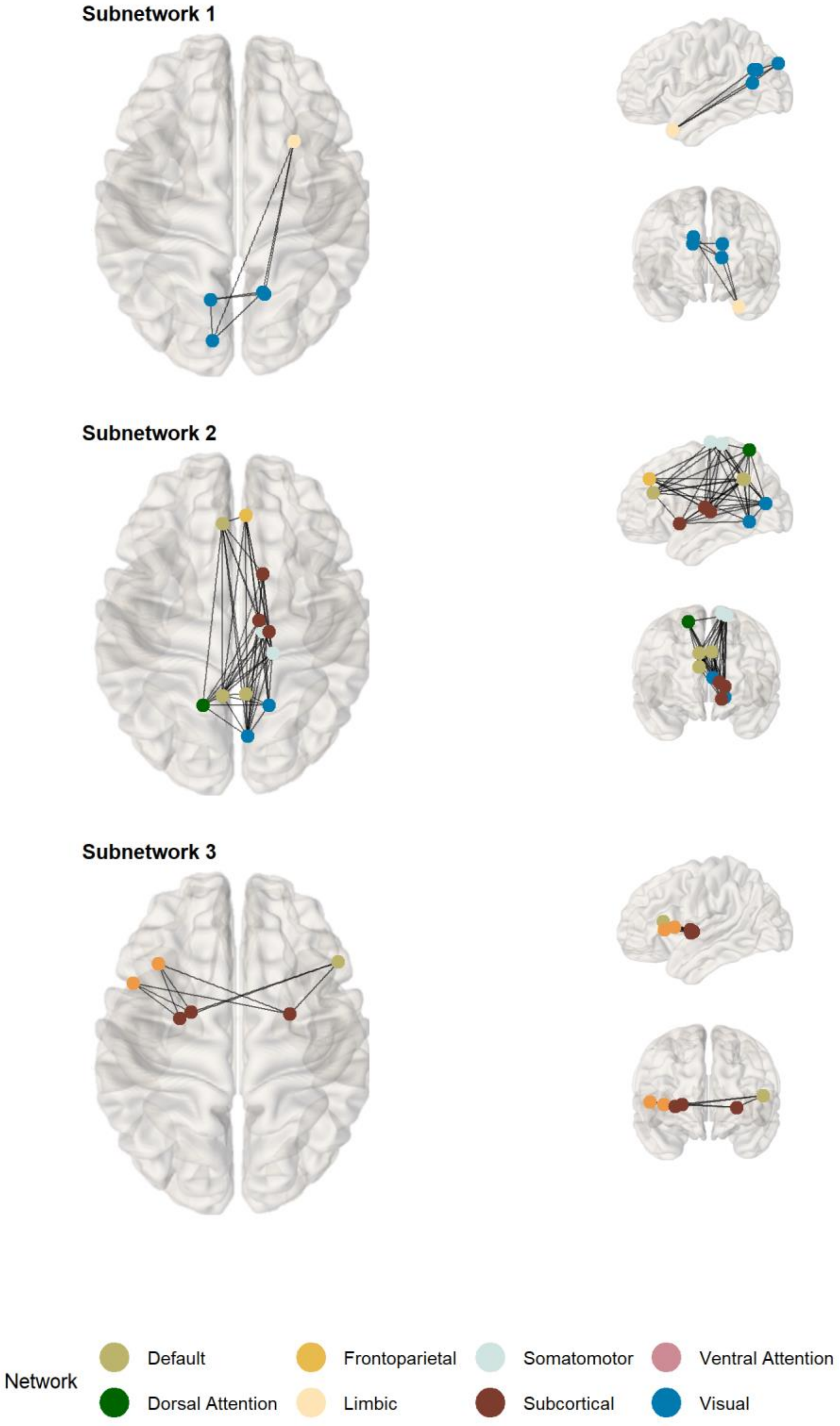


*Note.* Subgroup 3: most severe difficulties with hot executive skills & pragmatic communication.

**Figure S11**

*Subnetworks identified as significantly different from the comparison sample in Subgroup 1, Subgroup 2, and Subgroup 3 across several t-value thresholds. The figure demonstrates how the size of the identified subnetworks reduced with increasing t-value thresholds.*

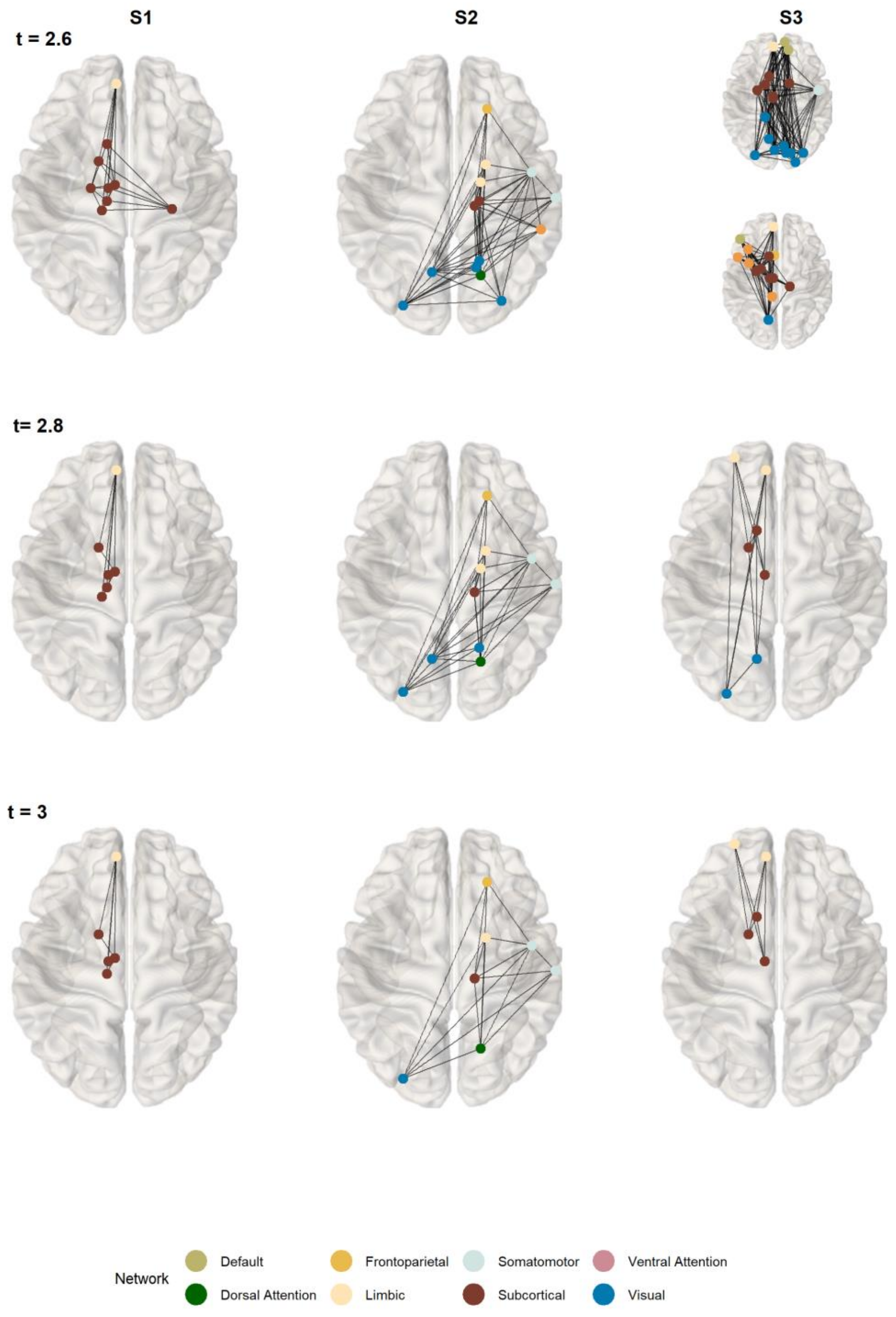


*Note.* Subgroup 1: most severe structural language difficulties; Subgroup 2: most severe *cool* executive difficulties; Subgroup 3: most severe difficulties with *hot* executive skills & pragmatic communication.

**Table S5**

*Edges within significant subnetwork identified for Subgroup 1*

| Regions of interest (Gyrus, Intrinsic connectivity network) | |
|---|---|
| Left medial area 11 (Orbital gyrus, Limbic) | Left nucleus accumbens (Basal ganglia, Subcortical) |
| Left medial area 11 (Orbital gyrus, Limbic) | Left medial pre-frontal thalamus (Thalamus, Subcortical) |
| Left medial area 11 (Orbital gyrus, Limbic) | Left occipital thalamus (Thalamus, Subcortical) |
| Left medial pre-frontal thalamus (Thalamus, Subcortical) | Left caudal temporal thalamus (Thalamus, Subcortical) |
| Left medial area 11 (Orbital gyrus, Limbic) | Left lateral pre-frontal thalamus (Thalamus, Subcortical) |

*Note.* Subgroup 1: most severe structural language difficulties.

**Table S6**

*Edges within significant subnetwork identified for Subgroup 2*

| Regions of interest (Gyrus, Intrinsic connectivity network) | |
|---|---|
| Right lateral area 11 (Orbital gyrus, Frontoparietal) | Right TE1.0/TE1.2 (Superior temporal Gyrus, Somatomotor) |
| Right TE1.0/TE1.2 (Superior temporal gyrus, Somatomotor) | Right caudal area 7 (Superior parietal lobule, Dorsal attention) |
| Right caudal area 22 (Superior temporal gyrus, Somatomotor) | Right caudal area 7 (Superior parietal lobule, Dorsal attention) |
| Right area 28/34 (EC entorhinal cortex) (Parahippocampal gyrus, Limbic) | Right caudal area 7 (Superior parietal lobule, Dorsal attention) |
| Right area TI [temporal agranular insular cortex] (Parahippocampal gyrus, Limbic) | Right caudal area 7 (Superior Parietal lobule, Dorsal attention) |
| Right caudal area 7 (Superior parietal lobule, Dorsal attention) | Left dorsomedial parietooccipital sulcus (Precuneus, Visual) |
| Right caudal area 7 (Superior parietal Lobule, Dorsal attention) | Left inferior occipital gyrus (Lateral occipital cortex, Visual) |
| Right rostral lingual gyrus (Medioventral occipital cortex, Visual) | Left inferior occipital gyrus (Lateral occipital cortex, Visual) |
| Right caudal area 7 (Superior parietal lobule, Dorsal attention) | Right posterior parietal thalamus (Thalamus, Subcortical) |

*Note.* Subgroup 2: most severe *cool* executive difficulties.

**Table S7**

*Edges within significant subnetwork identified for Subgroup 3*

| Regions of interest (Gyrus, Intrinsic connectivity network) | |
|---|---|
| Left lateral area10<br>(Middle frontal gyrus, Limbic) | Left middle occipital gyrus<br>(Lateral occipital cortex, Visual) |
| Left lateral area10<br>(Middle frontal gyrus, Limbic) | Left ventral caudate<br>(Basal ganglia, Subcortical) |
| Left medial area 11<br>(Orbital gyrus, Limbic) | Left ventral caudate<br>(Basal ganglia, Subcortical) |
| Left medial area 11<br>(Orbital gyrus, Limbic) | Left nucleus accumbens<br>(Basal ganglia, Subcortical) |
| Left dorsomedial parietooccipital sulcus<br>(Precuneus, Visual) | Left nucleus accumbens<br>(Basal ganglia, Subcortical) |
| Left lateral area10<br>(Middle frontal gyrus, Limbic) | Left rostral temporal thalamus<br>(Thalamus, Subcortical) |
| Left medial area 11<br>(Orbital gyrus, Limbic) | Left rostral temporal thalamus<br>(Thalamus, Subcortical) |
| Left ventral caudate<br>(Basal ganglia, Subcortical) | Left rostral temporal thalamus<br>(Thalamus, Subcortical) |

*Note.* Subgroup 3: most severe difficulties with *hot* executive skills & pragmatic communication.
